# Multi-ancestry polygenic risk scores for the prediction of type 2 diabetes and complications in diverse ancestries

**DOI:** 10.1101/2025.07.21.25331778

**Authors:** Alicia Huerta-Chagoya, Joohyun Kim, Ravi Mandla, Yingchang Lu, Ken Suzuki, Lauren E. Petty, Hong Kiat Ng, Jaewon Choi, Simon Lee, Madhusmita Rout, Kuang Lin, Linda S. Adair, Adebowale Adeyemo, Habibul Ahsan, Masato Akiyama, Ping An, Sonia S. Anand, Diane M. Becker, Alain G. Bertoni, Zheng Bian, Lawrence F. Bielak, John Blangero, Michael Boehnke, Erwin P. Bottinger, Donald W. Bowden, Fiona Bragg, Jennifer A. Brody, Thomas A. Buchanan, Brian E. Cade, Jin-Fang Chai, John C. Chambers, Giriraj R. Chandak, Li-Ching Chang, Kyong-Mi Chang, Miao-Li Chee, Chien-Hsiun Chen, Yuan-Tsong Chen, Zhengming Chen, Yii-Der I. Chen, Ji Chen, Guanjie Chen, Shyh-Huei Chen, Wei-Min Chen, Ching-Yu Cheng, Yoon Shin Cho, Hyeok Sun Choi, Lee-Ming Chuang, Miguel Cruz, Mary Cushman, Swapan K. Das, Ralph A. DeFronzo, H Janaka deSilva, Latchezar Dimitrov, Ayo P. Doumatey, Shufa Du, Qing Duan, Ravindranath Duggirala, Leslie S. Emery, James C. Engert, Daniel S. Evans, Michele K. Evans, Sarah Finer, Jose C. Florez, James S. Floyd, Myriam Fornage, Elizabeth G. Frankel, Barry I. Freedman, Lourdes García-García, Pauline Genter, Hertzel C. Gerstein, Mark O. Goodarzi, Penny Gordon-Larsen, Mariaelisa Graff, Myron Gross, Yu Guo, Xiuqing Guo, Yang Hai, Craig L. Hanis, MGeoffrey Hayes, Momoko Horikoshi, Annie-Green Howard, Sarah Hsu, Willa Hsueh, Wei Huang, Mengna Huang, Yi-Jen Hung, Mi Yeong Hwang, Chii-Min Hwu, Sahoko Ichihara, Michiya Igase, Eli Ipp, Mohammad T. Islam, Masato Isono, Hye-Mi Jang, Farzana Jasmine, Jost B. Jonas, Yoonjung Y. Joo, Edmond Kabagambe, Takashi Kadowaki, Yoichiro Kamatani, Fouad R. Kandeel, Sharon L.R. Kardia, Elizabeth W. Karlson, Anuradhani Kasturiratne, Norihiro Kato, Tomohiro Katsuya, Varinderpal Kaur, Takahisa Kawaguchi, Jacob M. Keaton, Abel N. Kho, Chiea-Chuen Khor, Muhammad Kibriya, Bong-Jo Kim, Woon-Puay Koh, Katsuhiko Kohara, Jaspal S. Kooner, Charles Kooperberg, Raymond J. Kreienkamp, Amel Lamri, Leslie A. Lange, Nanette R. Lee, Myung-Shik Lee, Jung-Jin Lee, Donna M. Lehman, Liming Li, Yun Li, Victor JY. Lim, Jianjun Liu, Yongmei Liu, Simin Liu, Jirong Long, Tin Louie, Xi Luo, Jun Lv, Julie A. Lynch, Shiro Maeda, Anubha Mahajan, Nisa M. Maruthur, Fumihiko Matsuda, Mark I. McCarthy, Roberta McKean-Cowdin, James B. Meigs, Iona Y. Millwood, Karen L. Mohlke, Ayesha A. Motala, Girish N. Nadkarni, Jerry L. Nadler, Masahiro Nakatochi, Mike A. Nalls, Uma Nayak, Aude Nicolas, Kari E. North, Darryl Nousome, Yukinori Okada, Ian Pan, James S. Pankow, Guillaume Paré, Jaehyun Park, Kyong Soo Park, Esteban J. Parra, Sanjay R. Patel, Mark A. Pereira, Patricia A. Peyser, Fraser J. Pirie, Michael Preuss, Michael A. Province, Bruce M. Psaty, Leslie J. Raffel, Laura M. Raffield, Laura J. Rasmussen-Torvik, Susan Redline, Alexander P. Reiner, Stephen S. Rich, Rebecca Rohde, Kathryn Roll, Rashedeh Roshani, Charles N. Rotimi, Charumathi Sabanayagam, Danish Saleheen, Kevin Sandow, Claudia Schurmann, Mohammad Shahriar, Douglas M. Shaw, Wayne H-H. Sheu, Jinxiu Shi, Xiao-Ou Shu, Megan M. Shuey, Moneeza K. Siddiqui, Jennifer A. Smith, Tamar Sofer, Cassandra N. Spracklen, Adrienne M. Stilp, Meng Sun, Yasuharu Tabara, E-Shyong Tai, Salman M. Tajuddin, Atsushi Takahashi, Fumihiko Takeuchi, Jingyi Tan, Kent D. Taylor, Katherine Taylor, Farook Thameem, Lin Tong, Fuu-Jen Tsai, Philip S. Tsao, Miriam S. Udler, Adan Valladares-Salgado, David A. van Heel, Rob M. vanDam, Rohit Varma, Maheak Vora, Niels Wacher-Rodarte, Ya-Xing Wang, Ellie Wheeler, Eric A. Whitsel, Ananda R. Wickremasinghe, Genevieve L. Wojcik, Tien Y. Wong, Jer-Yuarn Wu, Yong-Bing Xiang, Anny H. Xiang, Chittaranjan S. Yajnik, Ken Yamamoto, Toshimasa Yamauchi, Lisa R. Yanek, Jie Yao, Mitsuhiro Yokota, Canqing Yu, Jian-Min Yuan, Salim Yusuf, Eleftheria Zeggini, Liang Zhang, Weihua Zhang, Wei Zheng, Alan B. Zonderman, ENSA Genomics Consortium, Genes & Health Research Team, VA Million Veteran Program, Carlos A. Aguilar-Salinas, Clicerio González-Villalpando, Christopher A. Haiman, Young Jin Kim, Soo Heon Kwak, Aaron Leong, Ruth J.F. Loos, Andres Moreno-Estrada, Andrew P. Morris, Lorena Orozco, Jerome I. Rotter, Dharambir Sanghera, Teresa Tusie-Luna, Benjamin F. Voight, Marijana Vujkovic, Robin G. Walters, Tian Ge, Alisa K. Manning, Marie Loh, Jennifer E. Below, Xueling Sim, Josep M. Mercader, Maggie C.Y. Ng, D-PRISM Consortium

## Abstract

**Background:** Polygenic risk scores (PRSs) improve type 2 diabetes (T2D) prediction beyond clinical risk factors but perform poorly in non-European populations, where T2D burden is often higher, undermining their global clinical utility.

**Methods:** We conducted the largest global effort to date to harmonize T2D genome-wide association study (GWAS) meta-analyses across five ancestries—European (EUR), African/African American (AFR), Admixed American (AMR), South Asian (SAS), and East Asian (EAS)—including 360,000 T2D cases and 1·8 million controls (41% non-EUR). We constructed ancestry-specific and multi-ancestry PRSs in training datasets including 11,000 T2D cases and 32,000 controls, and validated their performance in independent datasets including 39,000 T2D cases and 126,000 controls of diverse ancestries. In the All of Us Research Program, we compared these PRSs to those from the Polygenic Score Catalog and assessed their ability to predict diabetes micro- and macrovascular complications.

**Findings:** Ancestry-specific PRSs showed limited prediction power for T2D in AFR, AMR, and SAS compared to EUR and EAS. In contrast, multi-ancestry PRSs, built using GWAS data from five ancestries, substantially improved T2D prediction across all ancestries. Compared to those in the interquartile range, individuals at the 97·5^th^ percentile of their PRSs had a 6-fold increased T2D risk in AMR, EAS, and EUR, and ≥3-fold in AFR and SAS. These PRSs were also associated with the development of microvascular complications and outperformed all previously reported PRSs for all ancestries.

**Interpretation:** We developed and extensively validated the most up-to-date T2D PRSs across diverse ancestry groups. These PRSs are publicly available to support further evaluation of their clinical utility in diverse ancestries.

## Introduction

Type 2 diabetes (T2D) represents one of the largest health problems of the 21st century, affecting 537 million people globally, and is predicted to increase to 783 million by 2045.^1^ Both genetic and environmental factors contribute to T2D susceptibility. Genome-wide association studies (GWAS) have identified 1,289 genetic signals associated with T2D in diverse ancestries.^2,3^ The aggregation of risk alleles in polygenic risk scores (PRSs) for T2D^2,4^ may provide insights into disease progression and prognosis, or help identify people at risk for prioritization of therapeutic or lifestyle intervention.^5^ Several initiatives are beginning to test their utility in clinical settings^6,7^, and an observational study in Electronic Health Record (EHR) data has shown that T2D PRSs provide data orthogonal to standard clinical risk factors and are therefore particularly valuable in identifying at-risk individuals among those perceived to be low risk based on standard clinical risk factors.^8^

T2D disproportionately affects people of ancestral genetic backgrounds other than European, with South Asian, African American, and Hispanic/Latin American populations having a higher prevalence of diabetes and related complications.^9^ Yet, most PRSs are based on GWAS, including predominantly individuals of European ancestry, and have poor performance in individuals of other ancestries, which may further exacerbate health disparities if PRS were to be deployed for disease prediction in ancestrally diverse populations.^2,10^

While efforts have been made to enhance PRS performance in diverse populations^11^, a comprehensive, standardized, and harmonized approach for the development and validation of PRSs across continental ancestries for T2D remains lacking. In particular, recent efforts have aimed at improving the transferability of PRS across diverse ancestries. First, the Polygenic Risk Methods in Diverse Populations (PRIMED) consortium and others have developed new methods, such as those incorporating GWAS and linkage disequilibrium (LD) data from multiple ancestries, showing improved prediction.^11,12^ Second, large-scale T2D GWAS data for populations other than European have become available, which may improve the power of PRS development in these populations. Third, the emergence of large-scale biobank data from around the world — such as the All of Us Research Program (AoU) — offers valuable opportunities to develop, train, and validate novel PRSs, as data from these participants has not yet been included in GWAS meta-analyses.^13,14^

As part of the Type 2 Diabetes Global Genomics Initiative (T2DGGI), we published the largest multi-ancestry T2D GWAS meta-analysis to date, based on 2·5 million individuals, including 428,452 with T2D, and enumerating 1,289 signals for T2D risk.^3^ Here, in collaboration with T2DGGI, we present results from the Diabetes Polygenic Risk Scores in Multiple ancestries (D-PRISM)^2,11^, an international consortium focusing on improving PRS prediction of different types of diabetes and progression across the lifespan in diverse ancestries. We leveraged these two consortia to aggregate extensive T2D GWAS data from five major continental ancestry groups, trained and validated PRS models in independent cohorts to evaluate their performance in predicting T2D and diabetes complications using a unified pipeline. We tested the optimal PRS model for each of the five ancestries and made them available to the community for downstream analyses, as well as future testing for clinical utility and implementation.

## Methods

An overview of the overall strategy is shown in Fig.1 and detailed in the supplementary methods. We leveraged T2D GWAS from selected cohorts participating in three large consortia: the Diabetes Meta-analysis of Trans-ethnic Association Studies (DIAMANTE)^15^, the Million Veteran Program (MVP)^16^, and FinnGen^17^. Cohorts were categorized by genetic similarity to one or more of the five ancestries available in the 1000 Genomes (1KG) project^18^, or the country of origin: African/African American (AFR), Admixed American (AMR), East Asian (EAS), European (EUR), and South Asian (SAS). While ancestry labels do not represent or account for the full continuum of human genetic diversity, they were necessary for statistical analyses where no individual-level data were available. We included 2,185,548 individuals (359,819 cases, 1,825,729 controls) across 125 T2D GWAS datasets to conduct ancestry-specific meta-analyses, with summary statistics from EUR and EAS ancestry groups representing 86% of the total sample size (EUR=68%, EAS=18%, AFR=6%, AMR=4%, and SAS=4%; Fig.1a, Supplementary Tables 1,2).

**Fig. 1.**
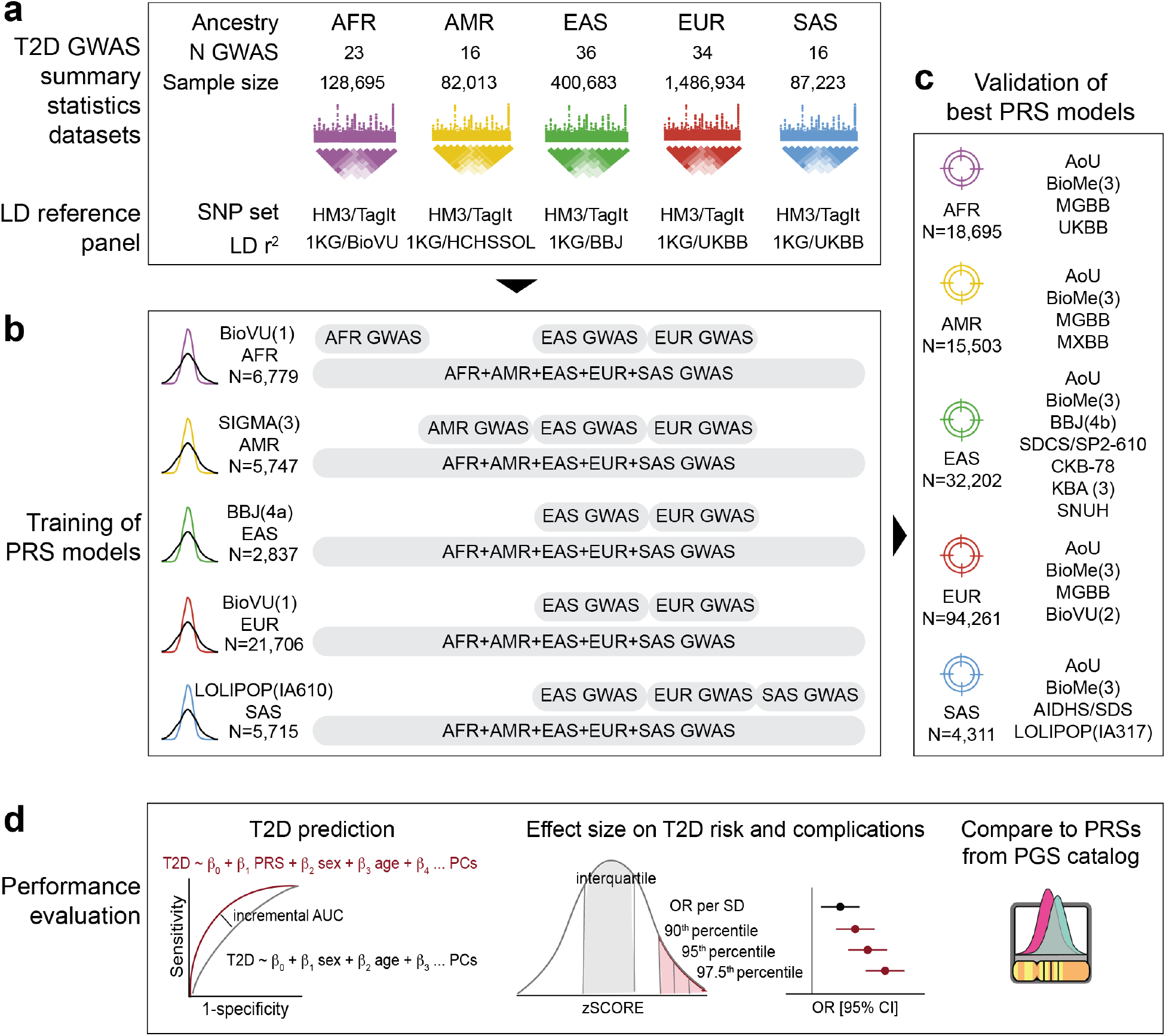
Overall analysis approach. **a,** Overview of the 125 T2D GWAS datasets, variant sets, and pairwise LD information used to generate five ancestry-specific T2D GWAS meta-analyses and 20 LD reference panels for PRSs training. **b,** Independent, ancestry-specific cohorts used to train the PRS models and select the optimal continuous shrinkage prior from five phi values (i.e., 0·01, 0·001, 1×10^−4^, 1×10^−5^, 1×10^−6^) based on predictive performance. Single-ancestry PRSs using GWAS summary statistics and LD panels matched to the validation ancestry or using data from the EAS or EUR ancestries. Multi-ancestry PRSs jointly modeled GWAS summary statistics and LD panels from all five ancestry groups. **c,** Set of 23 ancestry-specific and independent cohorts used to validate the 18 best-performing PRS. **d,** Evaluation of PRS predictive performance, including: i) incremental AUC, calculated as the difference between the AUC of the full model (PRS + covariates) and the model without the PRS, ii) proportion of variation in T2D status explained by the PRS, estimated using Nagelkerke’s r^2^, iii) odds ratio per standard deviation (OR per SD) of the PRS distribution or odds ratio (OR) comparing PRS distribution extremes relative to the interquartile range.

We leveraged cohorts not included in previous meta-analyses and, when necessary, conducted held-out meta-analyses to allow some cohorts to train and validate PRS. We aggregated data from a total of 42,784 individuals (10,992 cases, 31,792 controls) from five ancestries for training (tuning PRS construction parameters; Fig.1b) and 164,972 individuals (39,148 cases, 125,824 controls) for validation (testing PRS performance; Fig.1c). We used the five ancestry-specific T2D GWAS summary statistics to construct single-ancestry and multi-ancestry PRSs, using the PRS-CS (Continuous Shrinkage) ^19^ and PRS-CSx^20^ methods, respectively (Fig.1b). Both methods use LD panels from reference datasets to model the pairwise correlations between SNPs in GWAS summary statistics during PRS construction. The SNPs in the most widely used LD panels are based on the HapMap3 (HM3), which generally do not tag well in non-EUR populations and may miss ancestry-specific signals. In addition, the samples used to model LD structure are either from the 1KG project or the UK Biobank (UKBB). The former has a limited sample size, while the latter lacks sufficient representation of populations other than the EUR. To improve tagging, we developed new LD panels with an expanded set of SNPs generated using the Tag(ging) It(erative) of SNPs in multiple populations (TagIt) program^21^ and variants with ancestry-specific minor allele frequency (MAF) ≥0·01 across samples from the 1KG. We also use >8,000 in-house samples to compute pairwise LD in each ancestry, enabling more accurate modeling of the LD structure (Fig.1a).

We compared the impact of several input parameters (GWAS ancestry, two SNP sets, and two LD sources) on PRS performance in the training cohorts. We defined the best models as maximizing the incremental AUC (iAUC) for predicting prevalent T2D, comparing a full model, including the PRS, sex, age, and genetic principal components (PCs), to a model without the PRS (Fig.1d). We validated the best-performing models for association with T2D in at least four independent cohorts from each ancestry group (Fig.1c) and fitted secondary models with body mass index (BMI) as an additional covariate. We estimated the effect size of the PRSs as the odds ratio per standard deviation (OR per SD) unit of the PRS, and calculated OR for individuals at the 90^th^, 95^th^, and 97·5^th^ PRS percentiles compared with the interquartile range as the reference. In the AoU cohort, we also compared the association of our best-performing T2D PRSs with other published T2D PRSs from the Polygenic Score (PGS) catalog, and tested their association with common diabetes complications (Fig.1d).

## Results

### Development of new LD reference panels to improve variant coverage and LD estimation in diverse populations

Compared to the LD panels using the HM3 set of variants, our new TagIt-based LD panels increased the proportion of SNPs being tagged (r^2^≥0·8) up to 2 folds. The best improvement was 7 folds for SNPs with MAF 0·01-0·05 in AFR ancestry (Supplementary Table 7, Supplementary Fig.1). In all ancestry groups, the best-performing PRSs were those constructed using the expanded TagIt set of variants and/or the recomputed pairwise LD with large sample sizes (Supplementary Table 8, Supplementary Fig.2), suggesting that the prediction performance benefited from increasing SNP coverage, better LD modeling, or both.

### Matched single-ancestry PRS performance is positively correlated with the sample size of GWAS summary statistics

We first trained PRSs (Supplementary Tables 3,4) using GWAS summary statistics and four different LD panels, each matched to the ancestry of the validation cohorts (Supplementary Tables 5,6). When using a single-ancestry summary statistics, matching the GWAS and LD panel ancestries to that of the validation dataset resulted in the best prediction performance for the EUR (iAUC =0·07-0·14) and EAS (iAUC=0·02-0·16) ancestries. However, the prediction was poorer for AFR (iAUC=0·02-0·03), AMR (iAUC=0·02-0·04), and SAS (iAUC=0·02-0·04) (Fig.2a-e, Supplementary Table 9).

**Fig. 2.**
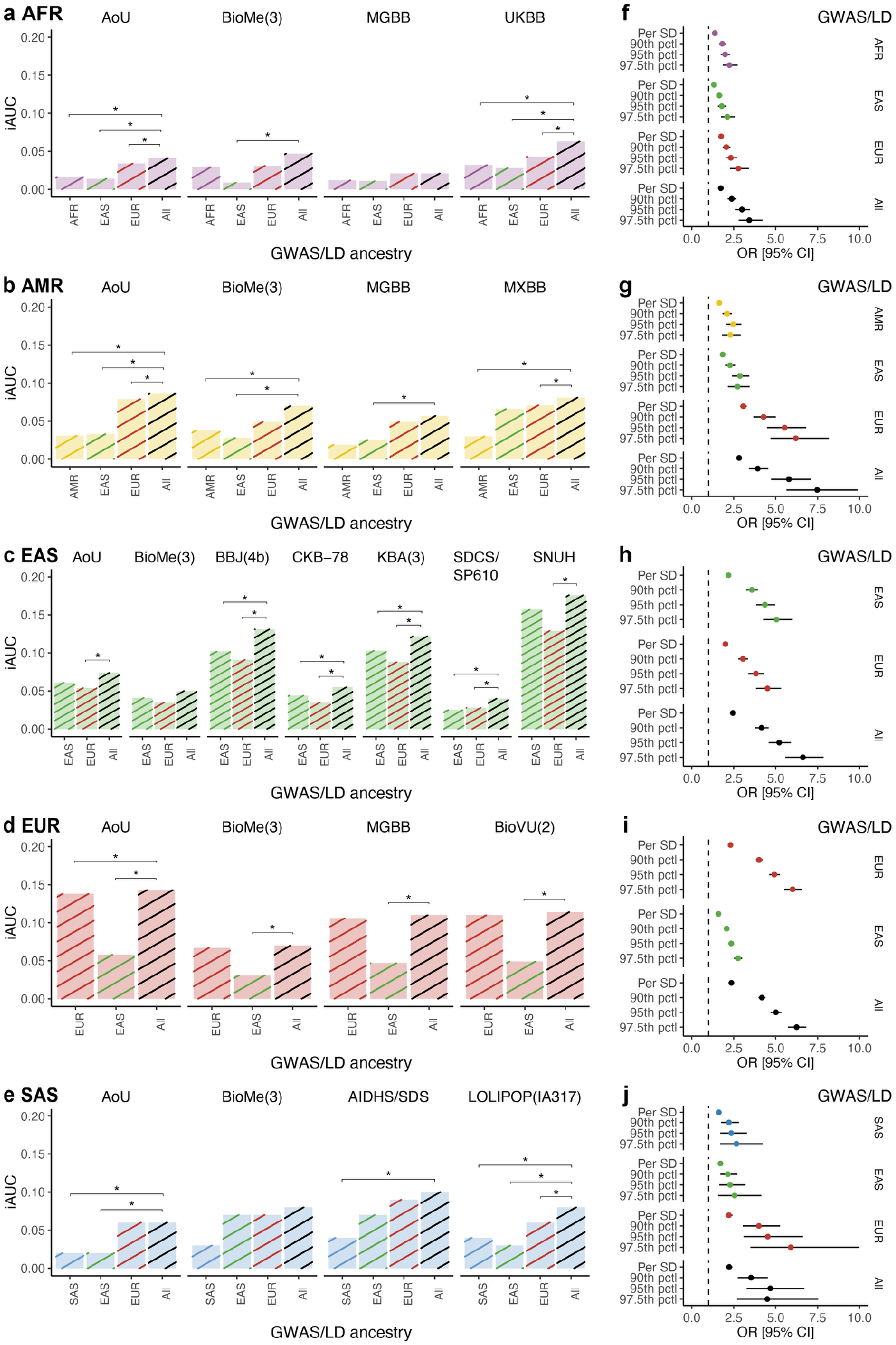
Performance of the T2D PRSs in the validation cohorts across ancestry groups. **a-e:** Incremental AUC (iAUC) of the T2D PRS in the validation cohorts across ancestry groups: **a,** AFR, **b,** AMR, **c,** EAS, **d,** EUR, **e,** SAS. For each ancestry, the best-performing single-ancestry and multi-ancestry PRSs were evaluated. Each bar represents a single cohort. Bar colors represent the ancestry group: purple for AFR, yellow for AMR, green for EAS, red for EUR, and blue for SAS. Line colors represent the ancestry of the T2D GWAS summary statistics and LD panels used to train the PRS, using the same color codes for single-ancestry PRSs, and black for multi-ancestry PRSs *De Long test p<0·05. **f-j:** Odds ratio from the meta-analysis of validation cohorts across ancestry groups: **f,** AFR, **g,** AMR, **h,** EAS, **i,** EUR, **j,** SAS. Points represent the odds ratio per standard deviation (OR per SD) of the PRS distribution or the odds ratio (OR) comparing different PRS distribution extremes relative to the interquartile range. Error bars show the 95% confidence intervals (95% CI). Point colors represent the ancestry of the T2D GWAS summary statistics and LD panels used to train the PRS. *De Long p<0.05.

The effect size of the PRS on T2D risk showed a similar pattern with larger OR per SD from the meta-analysis of the EUR (OR per SD [95% CI] = 2·31 [2·26-2·35]) and EAS (2·19 [2·12-2·27]) validation cohorts, compared to the AFR (1·38 [1·34-1·43]), AMR (1·64 [1·57-1·71]) and SAS (1·6 [1·5-1·74]). The EUR and EAS PRSs also showed better power to identify individuals at the highest T2D genetic risk than those trained for the other ancestries. For instance, individuals at the 90^th^ percentile of the respective PRS distribution had ∼4-fold increased risk of T2D in the EUR (OR [95% CI] = 4·01 [3·82-4·21]) and in the EAS (3·58 [3·24-3·94]) ancestries, compared to ∼2-fold increased risk for individuals in the AFR (1·82 [1·64-2·01], AMR (2·1 [1·84-2·39]) and SAS (2·22 [1·75-2·81]) ancestries (Fig.2f-j, Supplementary Table 9). We observed consistent results regardless of adjustment for BMI, indicating that the predictive performance of the PRSs was robust to the inclusion of this well-established T2D risk factor (Supplementary Table 10, Supplementary Fig.3).

### Using GWAS data from large non-matched ancestries improves PRS performance in ancestries with limited GWAS sample size

We then tested whether constructing PRS using GWAS summary statistics from ancestries with larger sample sizes, such as EUR and EAS, could improve T2D prediction in other ancestries with limited GWAS data, despite the larger ancestral differences between discovery and validation cohorts. Compared to the matched single-ancestry PRSs, those based on the EUR GWAS improved the T2D prediction in the AFR, AMR, and SAS validation cohorts but had lower predictions in the EAS.

A PRS based on EAS GWAS showed modestly improved T2D prediction in the AMR and SAS validation cohorts, but worse performance in the AFR and EUR (Fig.2a-e, Supplementary Table 9). The effect sizes of the PRSs were consistent with their prediction performance. The best-performing single-ancestry PRSs were the one derived from EUR GWAS for AFR (OR per SD (95% CI) = 1·75 [1·67-1·82]), AMR (3·07 [2·89-3·27]), SAS (2·21 [2·03-2·42]), and EUR (2·31 [2·26-2·35]) validation cohorts, while for EAS cohorts (2·19 [2·12-2·27]), the best-performing single-ancestry PRS was based on EAS GWAS (Fig.2f-j, Supplementary Table 9).

### Multi-ancestry PRSs show the best prediction performance in all ancestries

We further applied a multi-ancestry PRS method, PRS-CSx^20^, which jointly models ancestry-specific GWAS and LD panel data from multiple ancestries. This approach leverages the increased statistical power from GWAS of five continental ancestries, which are jointly modelled to maximize the power of variants that are present in all ancestries, while still incorporating the effects of variants that are specific or enriched in specific ancestries, even if their sample size is modest. Compared to the best single-ancestry PRSs, the prediction performance of the multi-ancestry PRSs was the highest across validation cohorts from all five ancestries (iAUC ranging from 0·02-0·06 in AFR, 0·06-0·09 in AMR, 0·04-0·17 in EAS, 0·07-0·14 in EUR, and 0·06-0·10 in SAS) (Fig.2a-e, Supplementary Table 9).

The multi-ancestry PRSs also had the highest effect sizes along with smaller confidence intervals across all ancestries (OR per SD [95% CI] = 1·73 [1·67-1·80] in AFR, 2·82 [2·67-2·97] in AMR, 2·45 [2·36-2·54] in EAS, 2·36 [2·32-2·41] in EUR, and 2·23 [2·05-2·42] in SAS). The improvement was particularly notable for individuals at the extremes of the PRS distributions. For instance, the individuals in the 97·5^th^ percentile of the multi-ancestry PRSs had 3 to 7-fold increased T2D risk compared to those in the interquartile range (OR [95% CI] = 3·43 [2·8-4·21] in AFR, 7·47 [5·64-9·89] in AMR, 6·62 [5·58-7·85] in EAS, 6·25 [5·72-6·82] in EUR and 4·50 [2·70-7·53] in SAS (Fig.2f-j, Supplementary Table 9).

### Multi-ancestry T2D PRSs outperform previously published T2D PRSs

We then leveraged the AoU cohort to compare this study’s best-performing multi-ancestry PRSs against the published T2D PRSs from the PGS catalog.^22^ We tested 55 out of 147 available PRS for T2D (accessed on October 07, 2024), after excluding pathway-specific PRSs or those for which the AoU cohort was used as a training dataset due to potential model overfitting. In all ancestry groups, our best-performing multi-ancestry PRSs showed better predictive performance than the previously published T2D PRSs from the PGS catalog. The differences with the best published PRSs from the PGS catalog were statistically significant for AFR, AMR, and EUR (De Long p<9×10^−4^, Bonferroni-corrected threshold=0·05/55 PRSs tested; iAUC for D-PRISM multi-ancestry PRS-CSx vs. best iAUC from PGS Catalog: 0·041 vs. 0·029 in AFR, 0·086 vs. 0·073 in AMR, 0·143 vs. 0·123 in EUR) (Fig.3a,b,d). The improvement was nominally significant for EAS (0·074 vs. 0·058, p=0·03) (Fig.3c), but not for SAS (p=0·58) (Fig.3e), likely due in part to their limited sample sizes in the AoU cohort for these two ancestries (Supplementary Table 11, Supplementary Fig.4-8).

**Fig. 3.**
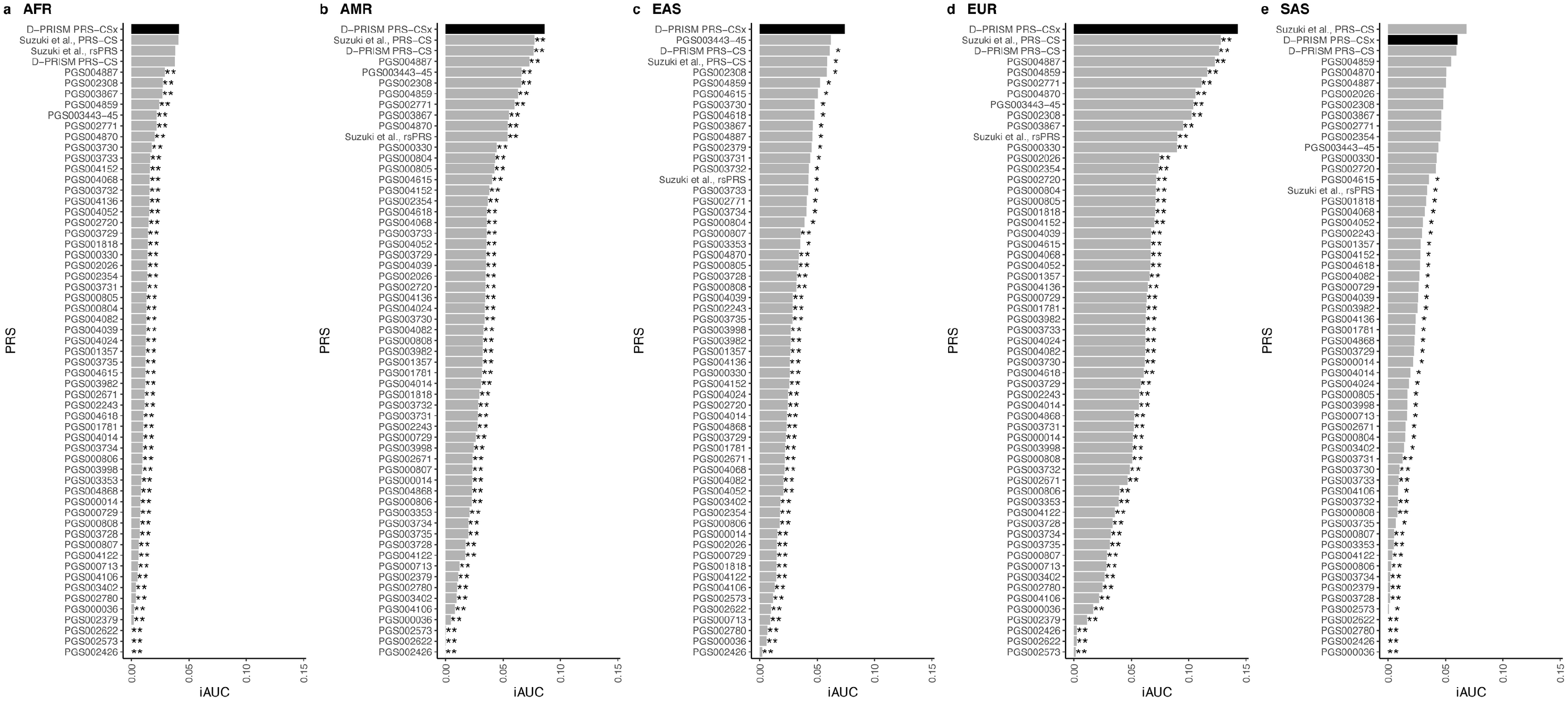
Performance of D-PRISM multi-ancestry PRSs compared to published T2D PRSs from the PGS Catalog and other sources in the All of Us cohort. **a-e:** Incremental AUC (iAUC) across ancestry groups: **a,** AFR, **b,** AMR, **c,** EAS, **d,** EUR, **e,** SAS. Black bars highlight this study’s multi-ancestry PRSs. *De Long p<0.05; **Bonferroni-corrected De Long p<9×10^−4^.

We also compared different strategies for PRS construction. Some studies leverage ancestry diversity using the inverse variance-weighted (IVW) meta-analysis results from multi-ancestry GWAS, typically applying a single LD panel — often of EUR ancestry— which may fail to model LD patterns and tag ancestry-specific variants accurately. In contrast, PRS-CSx jointly models GWAS from multiple ancestries while accounting for differences in allele frequencies and LD patterns across ancestries. To assess the value of PRS-CSx, we constructed a PRS-CS model using summary statistics from the IVW GWAS meta-analysis of the D-PRISM ancestry-specific GWAS summary statistics and tested its performance in the validation cohorts. In all ancestry groups, the T2D prediction performance of the multi-ancestry PRS-CSx was better than that of the PRS-CS models constructed using the IVW GWAS meta-analysis. The improvement was statistically significant in the AMR, EAS and EUR (iAUC for the multi-ancestry PRS-CSx vs. the multi-ancestry PRS-CS: 0·086 vs. 0·077 in AMR, 0·074 vs. 0·061 in EAS, and 0·143 vs. 0·126 in EUR, De Long p<0·05) (Fig.3b-d) but not significantly different for AFR and SAS (0·041 vs. 0·038 in AFR, p=0·07, 0·061 vs. 0·060 in SAS, p=0·91) (Fig.3a,e, Supplementary Table 11, Supplementary Figures 4-8).

**Fig. 4.**
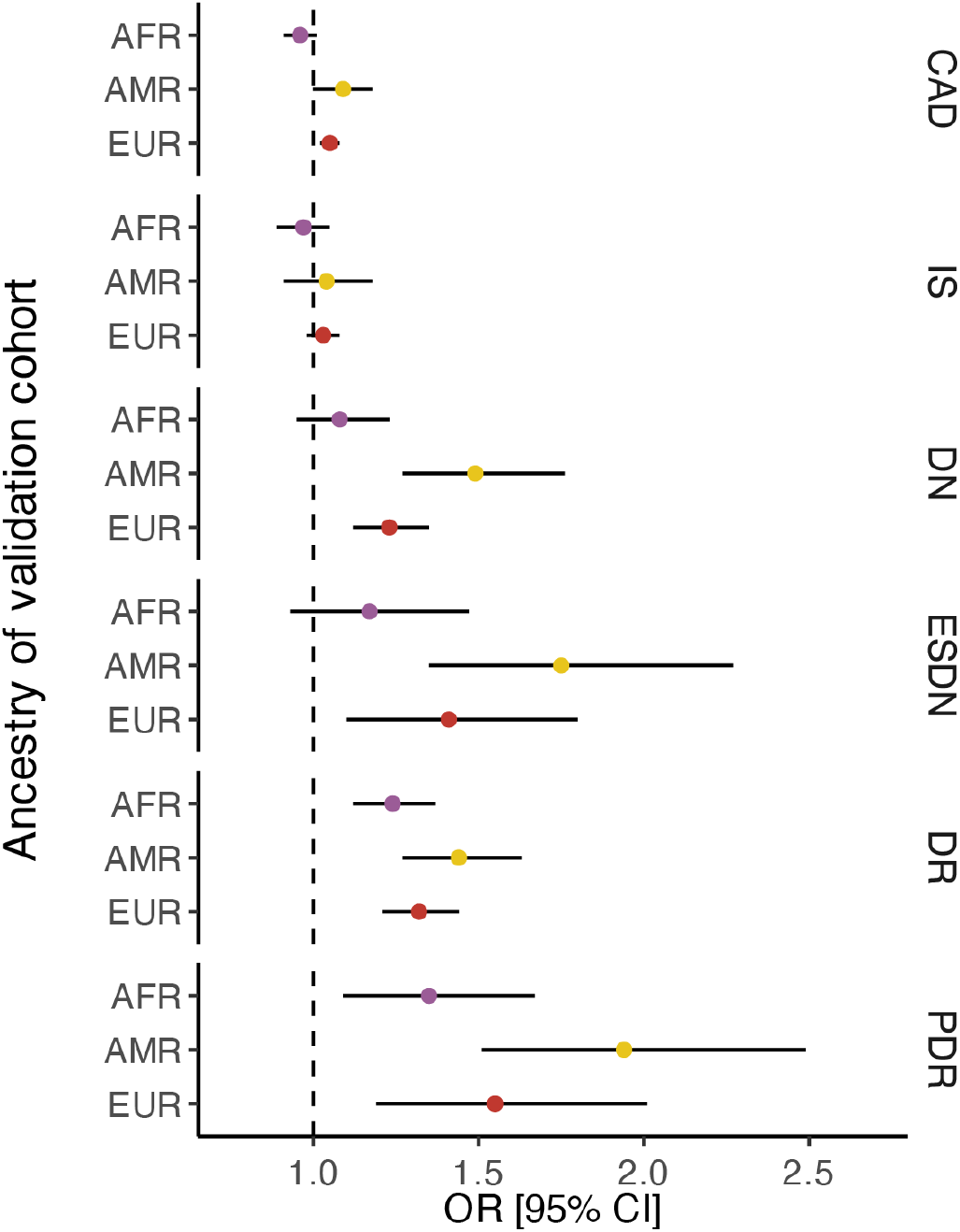
Association of D-PRISM multi-ancestry PRS with common complications of diabetes in the All of Us cohort. Odds ratio of this study’s multi-ancestry PRS for the following outcomes: CAD (cardiovascular disease), IS (ischemic stroke), DN (diabetic nephropathy), ESDN (end-stage diabetic nephropathy), DR (diabetic retinopathy), and PDR (proliferative diabetic retinopathy). Points represent the odds ratios per standard deviation (OR per SD) and are colored according to the genetic ancestry of the individuals tested: purple for AFR, yellow for AMR, and red for EUR. Error bars show the 95% confidence intervals (95% CI).

The largest multi-ancestry T2D GWAS meta-analysis to date, conducted by Suzuki et al.,^3^ includes a sample size 16% larger than our D-PRISM GWAS datasets (N=2,535,601, 428,452 cases, 2,107,149 controls), as we intentionally held out several cohorts for PRS training and validation in this study. Nevertheless, since the AoU cohort was not included in the discovery GWAS by Suzuki et al., we also evaluated the performance of a PRS-CS model built using those summary statistics. Despite a smaller sample size, our D-PRISM multi-ancestry PRS-CSx showed better prediction performance in the AMR, EUR, and EAS ancestry groups (iAUC for D-PRISM multi-ancestry PRS-CSx vs. Suzuki et al. PRS-CS: 0·086 vs. 0·077 in AMR, 0·074 vs. 0·059 in EAS, and 0·143 vs. 0·128 in EUR, De Long p<0·05) (Fig.3b-d). In the AFR and SAS ancestry groups, the D-PRISM multi-ancestry PRS-CSx yielded prediction performance compared to the Suzuki et al. PRS-CS model (0·041 vs. 0·041, p=0·82 in AFR, 0·061 vs. 0·068, p=0·34 in SAS) (Fig.3a,e).

In addition, our D-PRISM multi-ancestry PRS-CSx models showed consistently better prediction performance in four of the five ancestries (i.e., AMR, EAS, EUR, and SAS, p<0·05) (Fig.3b-e) compared to a multi-ancestry PRS that was restricted to the 1,289 genome-wide significant variants (rsPRS) identified in the discovery GWAS by Suzuki et al. We did not observe significant improvement in the AFR ancestry (Fig.3a, Supplementary Table 11, Supplementary Figs 4-8).

### Multi-ancestry T2D PRSs are associated with the risk of microvascular diabetes complications

T2D adversely affects the functioning of multiple organs, and the long-term complications accompanying the disease contribute the greatest morbidity for patients suffering from T2D.^23^ We leveraged the AoU cohort to assess if the best-performing multi-ancestry T2D PRSs may be helpful to identify individuals who are at high risk of developing diabetes associated macrovascular [i.e., cardiovascular disease (CAD), ischemic stroke (IS)] and microvascular complications [i.e., diabetic retinopathy (DR), proliferative diabetic retinopathy (PDR), diabetic nephropathy (DN), and end-stage diabetic nephropathy (ESDN)]. Given the sample size constraints of the AoU cohort, we only assessed the AFR, AMR, and EUR ancestries. We restricted microvascular complication analyses to individuals with T2D, as these outcomes are largely diabetes specific. For macrovascular complications, which also occur in those without T2D, we included all individuals and adjusted for T2D status in the models.

In each of the three ancestries, the D-PRISM multi-ancestry PRS-CSx was associated with increased risk of developing DR among individuals with T2D: OR per SD [95% CI] = 1·24 [1·12-1·37] in AFR, 1·44 [1·27-1·63] in AMR, 1·32 [1·21-1·44] in EUR. For the more severe form, PDR, the PRSs were also associated with increased risk in AFR, AMR, and EUR ancestries: 1·35 [1·09-1·67] in AFR, 1·94 [1·51-2·49] in AMR, 1·55 [1·19-2·01] in EUR. Additionally, the multi-ancestry PRSs predicted the risk for developing DN: 1·49 [1·27-1·76] in AMR and 1·23 [1·12-1·35] in EUR, as well as ESDN: 1·75 [1·35-2·27] in AMR and 1·41 [1·1-1·8] in EUR. It was also associated with increased risk of CAD in EUR only: 1·05 [1·02-1·08] (p<0·008, Bonferroni-corrected threshold from 0·05/6 diabetes complications tested) (Fig.4, Supplementary Table 12).

## Discussion

Identifying individuals with high T2D risk is essential to prioritize those who will benefit from lifestyle or therapeutic interventions to delay the disease or its complications.^24^ While clinical risk factors can identify individuals at risk, polygenic risk scores can be estimated at birth and can identify at risk individuals who may clinically be perceived at low risk, e.g., without a family history of T2D, young, lean, etc.^8^ Multiple efforts have been made to identify individuals at high genetic risk for T2D. More than a hundred PRS for T2D have been published^22^, yet they have poorer predictive performance in underrepresented populations. Several reasons influence the low transferability, including the overrepresentation of European populations in T2D GWAS and differential LD patterns between variants across ancestries. Importantly, populations in which PRSs are less predictive, including the AFR, AMR, and SAS, are disproportionately affected by diabetes and its complications, highlighting the risk of exacerbating health disparities by applying PRSs derived only from European genetic information.^11^ To improve the transferability and predictive accuracy of PRSs across diverse populations, including those underrepresented in GWAS efforts, various multi-ancestry PRS methods have been developed, while in parallel, several efforts have focused on expanding the representation of diverse ancestry populations in GWAS.^3,15,16,25,26^ Beyond GWAS discovery, multiple and sufficiently large sample sizes are also essential for training and validating PRSs, and rigorous data aggregation and harmonization following best practices are crucial to determine PRS accuracy.

In this study, we developed the most comprehensive PRSs for T2D across five continental ancestries and conducted extensive evaluations in at least four independent cohorts per ancestry. To accomplish this we leveraged nearly all available genetic datasets with T2D phenotype information to: i) maximize ancestry diversity by harmonizing 125 T2D GWAS datasets, including up to 2·2 million individuals; ii) enhance the representation and tagging of the genetic variants contributing to the PRS by generating new ancestry-specific LD reference panels, iii) train PRS models across ancestries and iv) thoroughly validate the PRSs in multiple independent harmonized cohorts and assess their association with diabetes-related complications.

As previously described^12,27^, we observed that PRSs have lower accuracy when the validation sample is genetically distant from the discovery GWAS sample. However, underrepresented populations in GWAS have limited power and imprecise variant effect size estimates. For this reason, PRSs derived from EUR GWAS continue to outperform those derived from ancestry-matched GWAS in AFR, AMR, and SAS ancestries, likely due to the EUR GWAS sample size being at least 11 times larger. In contrast, applying a EUR PRS yielded worse performance for EAS ancestry than using a matched-ancestry EAS GWAS, even though the latter was four times smaller than the EUR GWAS. This suggests that non-matched ancestry PRSs (i.e., PRSs based on GWAS from a different ancestry than the validation cohort) improve performance only when the GWAS sample size in the other ancestry is orders of magnitude larger than that of the matched-ancestry GWAS, which may lack power to capture ancestry-specific genetic effects. However, when power is sufficient, ancestry-matched GWAS can improve accuracy compared to larger cross-ancestry GWAS. The minimum ancestry-specific GWAS sample size for powerful PRS prediction may vary depending on the trait and ancestry and may require further investigation. Regardless, we observe that for all the ancestry groups, including EUR, which has the largest GWAS sample size^15^, the optimal PRS strategy is to combine PRSs from multiple ancestries, here using PRS-CSx. This approach can leverage the most accurate estimates from variants present in all ancestries while still accounting for population-specific or enriched variants, improving the overall prediction accuracy.

For all ancestries, the multi-ancestry PRSs showed the strongest associations with T2D risk among individuals at the extremes of the risk distribution. For instance, individuals of EUR, EAS, or AMR ancestry in the top 97·5^th^ percentile of the PRS distribution were associated with a seven-fold increased risk of developing T2D than those with average scores. Despite limited GWAS representation, the multi-ancestry PRS still shows the most predictive accuracy for individuals of AFR and SAS ancestry relative to the matched-ancestry PRS. However, overall performance remains lower than in other ancestry groups, with individuals in the 97·5^th^ percentile associated with three-fold and four-fold increased risk of T2D. These results underscore that, despite our best efforts, there is still a substantial gap among the performance of PRSs, particularly in these two ancestry groups.

Using data from the AoU cohort, we demonstrate that our multi-ancestry PRSs outperformed all previously available PRSs for T2D, likely because we used the largest and most diverse GWAS data and multi-ancestry-based methodology for PRS development. Additionally, we constructed our multi-ancestry PRSs using a standardized and rigorous approach and tested them extensively across diverse ancestry groups, supporting their broader applicability. For example, the multi-ancestry PRSs outperformed PRSs derived from the largest trans-ancestry meta-analysis. These findings underscore that, in addition to increasing representation in GWAS, leveraging methods that jointly model GWAS and LD panels across multiple ancestries can enhance the tagging of causal variants, thereby improving predictive performance. In contrast, standard methods that use multi-ancestry IVW GWAS meta-analysis results rely on a single LD reference panel and may fail to capture ancestry-specific LD patterns, limiting their ability to model genetic risk prediction accurately.

Previous studies have reported significant associations between T2D PRS, proliferative retinopathy, and end-stage diabetic nephropathy across and within ancestries.^15,16^ We confirm and extend previous findings by showing that multi-ancestry PRSs predict microvascular complications in T2D individuals of AFR, AMR, and EUR ancestries. Notably, our results reveal stronger associations and capture a broader range of diabetes complications severity than previously reported.

Our PRSs still face some limitations, mainly due to the lack of diversity in available genetic data. First, while constructing multi-ancestry PRS including GWAS from diverse ancestries—even those with smaller sample sizes— has proven beneficial to capture ancestry-specific effects, the SNP effect sizes are still strongly influenced by the largest European cohorts. Second, we acknowledge that using discrete population categories or restricting the analyses to groups defined by genetic similarity is sub-optimal, particularly in highly admixed populations where such groupings do not fully capture heterogeneous ancestry. While our current approach partially addresses these challenges, it remains limited by the need to group individuals into discrete ancestries, rather than a continuum. A critical next step will be adopting continuous genetic ancestry methods, which will require broader data access and new analytical strategies.

Despite criticism regarding PRSs’ limited added value to clinical risk factors, it has been shown that incorporating PRSs can help identify high-risk individuals among people who are clinically perceived as low risk, such as the young, the lean, or those with sparse clinical data^8^, and that PRSs improve prediction over family history alone^28^. Ongoing randomized controlled trials are starting to evaluate the impact of PRS implementation into adult primary care as an additional risk factor for the primary care provider to consider.^7,29^ Importantly, for any PRS-informed interventions to be viable, a state-of-the-art T2D PRS is a necessary first step. Among the characteristics to prioritize when using a PRS are its generalizability, portability, and implementation feasibility. For instance, Lennon *et al*.^6^ prioritized a multi-ancestry T2D PRS^30^ trained using EUR, EAS, and AFR GWAS data, and proposed a high-risk cut-off of 2%. Compared to the rest of the non-high-risk population, this PRS showed ORs ranging from 4·44 [3·60-5·49] in EUR to a maximum of 2·35 [1·54-3·60] in AFR cohorts. Notably, our multi-ancestry PRSs consistently improved the prediction performance of T2D irrespective of the individual’s continental ancestry over any previously existing PRS. Compared to people with average genetic risk, those above the high genetic risk cut-off (2·5%) have ORs ranging from 3·43 to 7·47 across ancestries, compared to the interquartile group. We therefore propose using the T2D PRS models presented here for future clinical applications as they show improved predictive ability across all ancestries.

In summary, this study addresses a critical gap by delivering the most comprehensive and rigorously tested set of multi-ancestry PRSs for T2D. These scores improve risk prediction across diverse populations and enhance the identification of individuals at high genetic risk of developing T2D and microvascular complications, including mild and severe forms. By making the PRS weights publicly available, we provide a valuable resource for researchers and clinicians seeking to advance genetic risk stratification and develop prevention strategies for T2D.

## Contributors

J.M.M. and M.C.Y.N. conceived the study. A.H-C. and J.K. led the analyses. R.M., Y.L., K.S., L.E.P., H.K.N., J.C., S.L., M.R., and K.L. performed the training and/or validation of the PRS analyses. Y.J.K., R.G.W., D.S., S.H.K., and R.J.F.L. supervised the training and/or validation of the PRS analyses. T.G., A.K.M., M.L., J.E.B., X.S., J.M.M and M.C.Y.N. jointly directed this work. The rest of the authors provided data for the different stages of the analyses. A.H.-C., J.K., J.M.M., and M.C.Y.N. wrote the first draft of the manuscript. All authors contributed to interpreting the data, and they read, revised, and approved the final manuscript.

## Data sharing

Individual participant data is not available because they are subject to data protection laws and restrictions imposed by the ethics committee to ensure study participants’ privacy. The study protocol and the individual methods are included in the methods section. The developed PRS weights are available without restrictions through the PGS catalog (https://www.pgscatalog.org).

## Data Availability

The study protocol and the individual methods are included in the methods section. The developed PRS weights are available without restrictions through the PGS catalog (https://www.pgscatalog.org).

## Acknowledgments

This work is supported by National Human Genome Research Institute (NHGRI) of the National Institutes of Health (NIH) U01HG011723. A.H.-C. is supported by the American Diabetes Association (ADA) grant 11-23-PDF-35. Y.L. is supported by R56HL150186, R01HL158884, and R01DK135938. M.O.G. was supported in part by NIH grants from the National Institute of Diabetes and Digestive and Kidney Disease (NIDDK) (P30-DK063491) and from the National Center for Advancing Translational Sciences (NCATS) (UL1TR001420, UL1TR001881) and the Eris M. Field Chair in Diabetes Research. S.S.R. is supported by NHGRI U01HG011723. M.A.N. research was supported in part by the Intramural Research Program of the NIH, National Institute on Aging (NIA), NIH, Department of Health and Human Services; project number ZO1 AG000535, as well as the National Institute of Neurological Disorders and Stroke (NINDS). This work utilized the computational resources of the NIH HPC Biowulf cluster. (http://hpc.nih.gov).

A.L. is supported by grant 2020096 from the Doris Duke Foundation, the ADA grant 7-22-ICTSPM-23, and NHGRI U01HG011723. A.P.M. acknowledges support from the NIHR Manchester Biomedical Research Centre (NIHR203308). C.N.S. was supported by the NIH (R01DK118011; R01DK136671) and the ADA (11-22-JDFPM-06). J.B.M. is supported by grant UMDK078616. B.F.V. is grateful for support from the NIDDK DK138521 and DK126194. A.K.M. is supported by grant UMDK078616. J.M.M. is supported by ADA grant #11-22-ICTSPM-16 and by NHGRI U01HG011723, by the NIDDK under Award Number R01DK137993 and U01 DK140757, AMP CMD award from RFP 6 from the Foundation for the NIH, and a Medical University of Bialystok (MUB) grant from the Ministry of Science and Higher Education (Poland). This work is supported by the Novo Nordisk Foundation (NNF21SA0072102). M.C.Y.N. is supported by U01HG011723, R01DK066358 and U01DK105556.

## References

1 Dianna J. Magliano, Edward J. Boyko, IDF Diabetes Atlas 10th edition scientific committee. IDF DIABETES ATLAS. 2021.

2 Mercader JM, Ng MCY, Manning AK, Rich SS. Predicting diabetes risk in diverse populations: what next? Lancet Diabetes Endocrinol 2021; 9: 808–10.

3 Suzuki K, Hatzikotoulas K, Southam L, et al. Genetic drivers of heterogeneity in type 2 diabetes pathophysiology. Nature 2024; 627: 347–57.

4 Udler MS, McCarthy MI, Florez JC, Mahajan A. Genetic Risk Scores for Diabetes Diagnosis and Precision Medicine. Endocr Rev 2019; 40: 1500–20.

5 Diabetes Prevention Program Research Group, Knowler WC, Fowler SE, et al. 10-year follow-up of diabetes incidence and weight loss in the Diabetes Prevention Program Outcomes Study. Lancet 2009; 374: 1677–86.

6 Lennon NJ, Kottyan LC, Kachulis C, et al. Selection, optimization and validation of ten chronic disease polygenic risk scores for clinical implementation in diverse US populations. Nat Med 2024; 30: 480–7.

7 Vassy JL, Brunette CA, Lebo MS, et al. The GenoVA study: Equitable implementation of a pragmatic randomized trial of polygenic-risk scoring in primary care. Am J Hum Genet 2023; 110: 1841–52.

8 Mandla R, Schroeder P, Porneala B, et al. Polygenic scores for longitudinal prediction of incident type 2 diabetes in an ancestrally and medically diverse primary care physician network: a patient cohort study. Genome Med 2024; 16: 63.

9 Fortmann AL, Savin KL, Clark TL, Philis-Tsimikas A, Gallo LC. Innovative Diabetes Interventions in the U.S. Hispanic Population. Diabetes Spectr 2019; 32: 295–301.

10 Martin AR, Kanai M, Kamatani Y, Okada Y, Neale BM, Daly MJ. Clinical use of current polygenic risk scores may exacerbate health disparities. Nat Genet 2019; 51: 584–91.

11 Kullo IJ, Conomos MP, Nelson SC, et al. The PRIMED Consortium: Reducing disparities in polygenic risk assessment. Am J Hum Genet 2024; 111: 2594–606.

12 Kachuri L, Chatterjee N, Hirbo J, et al. Principles and methods for transferring polygenic risk scores across global populations. Nat Rev Genet 2024; 25: 8–25.

13 Kidenya BR, Mboowa G. Inclusiveness of the All of Us Research Program improves polygenic risk scores and fosters genomic medicine for all. Communications medicine 2024; 4: 227.

14 All of Us Research Program Genomics Investigators. Genomic data in the All of Us Research Program. Nature 2024; 627: 340–6.

15 Mahajan A, Spracklen CN, Zhang W, et al. Multi-ancestry genetic study of type 2 diabetes highlights the power of diverse populations for discovery and translation. Nat Genet 2022; 54: 560–72.

16 Vujkovic M, Keaton JM, Lynch JA, et al. Discovery of 318 new risk loci for type 2 diabetes and related vascular outcomes among 1.4 million participants in a multi-ancestry meta-analysis. Nat Genet 2020; 52: 680–91.

17 Kurki MI, Karjalainen J, Palta P, et al. FinnGen provides genetic insights from a well-phenotyped isolated population. Nature 2023; 613: 508–18.

18 1000 Genomes Project Consortium, Auton A, Brooks LD, et al. A global reference for human genetic variation. Nature 2015; 526: 68–74.

19 Ge T, Chen C-Y, Ni Y, Feng Y-CA, Smoller JW. Polygenic prediction via Bayesian regression and continuous shrinkage priors. Nat Commun 2019; 10: 1776.

20 Ruan Y, Lin Y-F, Feng Y-CA, et al. Improving polygenic prediction in ancestrally diverse populations. Nat Genet 2022; 54: 573–80.

21 Wojcik GL, Fuchsberger C, Taliun D, et al. Imputation-Aware Tag SNP Selection To Improve Power for Large-Scale, Multi-ethnic Association Studies. G3 (Bethesda) 2018; 8: 3255–67.

22 Lambert SA, Wingfield B, Gibson JT, et al. Enhancing the Polygenic Score Catalog with tools for score calculation and ancestry normalization. Nat Genet 2024; 56: 1989–94.

23 Farmaki P, Damaskos C, Garmpis N, Garmpi A, Savvanis S, Diamantis E. Complications of the Type 2 Diabetes Mellitus. Curr Cardiol Rev 2020; 16: 249–51.

24 Crandall JP, Knowler WC, Kahn SE, et al. The prevention of type 2 diabetes. Nat Clin Pract Endocrinol Metab 2008; 4: 382–93.

25 Loh M, Zhang W, Ng HK, et al. Identification of genetic effects underlying type 2 diabetes in South Asian and European populations. Commun Biol 2022; 5: 329.

26 Spracklen CN, Horikoshi M, Kim YJ, et al. Identification of type 2 diabetes loci in 433,540 East Asian individuals. Nature 2020; 582: 240–5.

27 Ding Y, Hou K, Xu Z, et al. Polygenic scoring accuracy varies across the genetic ancestry continuum. Nature 2023; 618: 774–81.

28 Drzymalla E, Raffield L, Kolor K, et al. Additive Value of Polygenic Risk Score to Family History for Type 2 Diabetes Prediction: Results From the All of Us Research Database. Diabetes Care 2025; 48: 212–9.

29 Linder JE, Allworth A, Bland HT, et al. Returning integrated genomic risk and clinical recommendations: The eMERGE study. Genet Med 2023; 25: 100006.

30 Ge T, Irvin MR, Patki A, et al. Development and validation of a trans-ancestry polygenic risk score for type 2 diabetes in diverse populations. Genome Med 2022; 14: 70.

