## Supplementary material for "Multi-ancestry polygenic risk scores for the prediction of type 2 diabetes and complications in diverse ancestries"

**Supplementary methods**

Type 2 diabetes GWAS meta-analyses used for the construction of the PRSs

We leveraged the T2D GWAS summary statistics from a subset of cohorts participating in three large Consortia: Diabetes Meta-analysis of Trans-ethnic Association Studies (DIAMANTE)[1], the Million Veteran Program (MVP)[2], and the FinnGen Study.[3] An independent subset of cohorts was used for the development and validation of the PRSs. If a cohort was multi-ancestry, each individual was categorized by genetic similarity to one or more of the five ancestries available in the 1000 Genomes (1KG) Project[4] and/or the Human Genome Diversity Project[5] as reference panels: African/African American (AFR), Admixed American (AMR), East Asian (EAS), European (EUR), and South Asian (SAS). For single-ancestry cohorts, the grouping was based on the country of recruitment. We included 2,185,548 individuals (359,819 T2D cases and 1,825,729 controls) across 125 T2D GWAS to conduct ancestry-specific meta-analyses (Fig.1a, Supplementary Tables 1,2).

Each GWAS tested the association of the genetic variants with T2D adjusted for age, sex, the top genetic principal components (PCs), and cohort-specific covariates. We performed an inverse variance weighted (IVW) fixed-effect meta-analysis for each ancestry group with the METAL software.[6] We then applied quality control to keep biallelic, nonpalindromic SNPs in at least half of the effective sample size with a minor allele frequency (MAF) ≥0·01. For each ancestry-specific T2D GWAS meta-analysis, we intended to include most cohorts to maximize the sample size of the summary statistics for constructing PRSs while leaving out sufficient cohorts for each ancestry to be used for training and validation.

Cohorts for the training and validation of the PRSs

We trained the PRSs in one cohort per ancestry group and validated them in at least four validation cohorts per ancestry. All training and validation cohorts included unrelated individuals and were independent of those included in the GWAS summary statistics to avoid overfitting (Fig.1b-c, Supplementary Tables 3-6). Except for the AoU cohort, for which whole genome sequencing is available, the genotyping of the other cohorts was chip array-based. The genotypes were imputed to the 1KG[4] or the TOPMed r2[7,8] reference panels using the Michigan Imputation server.[9] We applied a separate post-imputation quality control in each cohort and ancestry to keep biallelic nonpalindromic SNPs with an imputation quality of $r^2$≥0·8 and MAF≥0·005. We excluded the variants not included in the LD reference panels, as explained below, or variants that showed an allelic frequency discordance≥0·2 compared to the 1KG ancestry-specific allelic frequency.

LD reference panels for the construction of the PRSs

To account for the correlation between variants, we constructed customized ancestry-specific LD reference panels using the same scripts used in PRS-CS[10] and PRS-CSx[11] tools. We built four new sets of ancestry-specific LD reference panels using the HapMap3 (HM3) set of variants, similar to the official panel (https://github.com/getian107/PRScsx), or an expanded 1KG set of variants, along with pairwise LD from the 1KG or in-house samples.

First, we identified ancestry-specific LD blocks using LDetect.[12] For each of the five ancestry groups in the 1KG dataset (https://mathgen.stats.ox.ac.uk/impute/impute_v2.html#reference), we selected common SNPs with MAF≥0·01 to generate a covariance matrix of variants based on the LD $r^2$ calculated in PLINK v1·9[13] and derived the boundaries of LD blocks. Second, we generated two sets of reference SNPs. One was based on the HM3 set of variants (https://www.sanger.ac.uk/data/hapmap-3/), similar to the official PRS-CS[10]/PRS-CSx[11] HM3 version. Another expanded set of variants based on 1KG (https://mathgen.stats.ox.ac.uk/impute/impute_v2.html#reference) was selected using the Tag(ging) It(erative) of SNPs in multiple populations (TagIt) program.[14] TagIt allows the selection of tag SNPs by leveraging genetic information from multiple diverse ancestries to maximize cross-population coverage. We only included non-palindromic SNPs with MAF≥0·01 in at least one ancestry group for both the HM3 and TagIt SNP lists. Third, the two sets of SNPs were extracted in each ancestry for individuals from the 1KG dataset (347 to 661 individuals per ancestry) and from the larger imputed in-house datasets (including around 10,000 individuals per ancestry). SNPs with low imputation quality ($r^2$<0·8) were further excluded in the in-house datasets. Last, we calculated the variants' pairwise LD ($r^2$) using PLINK v1·9 to generate ancestry-specific LD reference panels. In total, for each ancestry, we built four different LD reference panels and used them to construct the PRSs, combining two sets of variants (i.e., HM3 and TagIt) and two sources of LD information (i.e., 1KG and in-house samples) (Supplementary Table 7).

Training of the PRSs

We used the PRS-CS[10] Bayesian polygenic method to construct single-ancestry PRSs. For each ancestry group, we leveraged the T2D GWAS summary statistics and LD reference panels matching the ancestry of the validation cohort (e.g., a PRS trained using the AMR GWAS and AMR LD reference panel to be validated in AMR cohorts). When applicable, we also modeled non-matched single-ancestry PRSs (e.g., a PRS trained using the EUR GWAS and EUR LD reference panel to be evaluated in AMR cohorts). The PRS-CS method returns a single-ancestry posterior variant effect size.

We then used PRS-CSx[11] to construct multi-ancestry PRSs. Instead of meta-analyzing the ancestry-specific GWAS summary statistics, PRS-CSx jointly models the GWAS summary statistics along with their matching LD reference panels, using a shared continuous shrinkage prior, to generate ancestry-specific variant posterior effect sizes in a coupled manner that leverages cross-population genetic architecture. We used these ancestry-specific effect sizes to compute standardized ancestry-specific z-scores, which were then combined in a linear regression model to derive the multi-ancestry posterior variant effect size as follows:

$$y = PRS_{\theta,AFR} + PRS_{\theta,AMR} + PRS_{\theta,EAS} + PRS_{\theta,EUR} + PRS_{\theta,SAS}$$

Where y is the T2D status, and $PRS_{\theta,\text{ ancestry group}}$ is the standardized PRS for a given shrinkage prior and ancestry.

For both PRS-CS and PRS-CSx, we used the training cohorts to select the optimal continuous shrinkage prior from five phi values (i.e., 0·01, 0·001, $1x10^{-4}$, $1x10^{-5}$, $1x10^{-6}$) based on predictive performance. In total, we constructed 80 PRSs for each of the AFR, AMR, and SAS ancestry groups (i.e., 1 matched-ancestry PRS, 2 non-matched ancestry PRSs, and 1 multi-ancestry PRS × 4 LD panels × 5 phi values = 80 models) and 60 PRSs for each of the EAS and EUR ancestry groups (i.e., 1 matched-ancestry PRS, 1 non-matched ancestry PRS, and 1 multi-ancestry PRS × 4 LD panels × 5 phi values = 60 models), resulting in 360 PRS models overall.

To test the predictive performance, we applied the posterior variant effect sizes for each PRS model to calculate the individual scores in each of the five training cohorts using the --score function in PLINK v1·9.[9] We standardized them to have a mean of zero and unit variance. Then, we fitted two logistic regression models and calculated the area under the receiver operator characteristic curve (AUC) using the "pROC" package[15] in R. One model included the explanatory variables sex, age, and genetic PCs, and a second full model also included the standardized PRS. We also fitted logistic regression models adjusted for BMI.

We calculated the incremental AUC (iAUC) by subtracting the AUC of the model without the PRS from the AUC of the full model. We defined the best-trained PRS models as those with the continuous shrinkage prior and LD panel that maximized the iAUC. After the training step, we ended up with four best-trained PRS models for the AFR, AMR, and SAS ancestries (i.e., one matched-ancestry PRS, one EUR non-matched ancestry PRS, one EAS non-matched ancestry PRS, and one multi-ancestry PRS), 3 best-trained PRS models for the EUR ancestry (i.e., one single matched-ancestry PRS, one EAS non-matched ancestry PRS, and one multi-ancestry PRS), and 3 best-trained PRS models for the EAS ancestry (i.e., one single matched-ancestry PRS, one EUR non-matched ancestry PRS, and one multi-ancestry PRS) (Supplementary Table 8).

Validation of the PRSs

To validate each of the 18 best-trained PRSs, we applied the posterior variant effect sizes in a second set of unrelated samples and independent cohorts from each ancestry group. We calculated the individual scores in each validation cohort using the --score function in PLINK 1·9[13] and standardized them to have a mean of zero and unit variance. For the multi-ancestry PRS, we combined ancestry-specific standardized scores weighted for the trained ancestry-specific effect sizes as follows:

$$y = \beta_{\theta,AFR}\, PRS_{\theta,AFR} + \beta_{\theta,AMR}\, PRS_{\theta,AMR} + \beta_{\theta,EAS}\, PRS_{\theta,EAS} + \beta_{\theta,EUR}\, PRS_{\theta,EUR} + \beta_{\theta,SAS}\, PRS_{\theta,SAS}$$

Where y is T2D status, $\beta_{\theta,\text{ ancestry group}}$ is the regression coefficient for a given shrinkage prior and ancestry in the training cohort, and $PRS_{\theta,\text{ ancestry group}}$ is the standardized PRS for a given shrinkage prior and ancestry.

To test the predictive performance of the PRS, we calculated i) the iAUC as explained above, ii) the proportion of the variation in the T2D status explained by the PRS using Nagelkerke's $r^2$, iii) the odds ratio per standard deviation (OR per SD) change in the PRS, and iv) the discrimination capacity at the extremes of the PRS distribution by identifying the individuals at the top 97·5th percentile, 95th percentile and 90th percentile for comparison with the interquartile range.

We applied the DeLong test to statistically assess the difference between AUCs of the single ancestry vs. multi-ancestry PRSs. We combined the PRS estimates across validation cohorts using fixed-effects meta-analyses by ancestry, weighting each cohort's beta coefficient by the inverse of its variance, using the "metafor" package[16] in R (Supplementary Tables 9,10).

Comparison of the best-performing multi-ancestry PRSs to other publishedT2D PRS

Multiple efforts have been made to improve the portability of PRS to apply to individuals from diverse ancestries. Until the preparation of this work (revised on October 07, 2024), 147 PRSs for T2D had been constructed and made publicly available through the PGS catalog.[17] To compare the performance of our best-trained PRS models using PRS-CSx (which we refer to as 'D-PRISM multi-ancestry PRS-CSx' model), we selected 55 PRSs from the PGS catalog that i) were trained for the overall T2D trait and not for any specific subtype of the disease, ii) were trained considering all types of genetic variants and not any specific set of variants for specific biological mechanisms, and iii) were constructed and trained by leveraging genetic information from cohorts other than the AoU, which we used as a validation cohort in this study.

We downloaded the PRSs and extracted the variants from the AoU cohort[18] (release v7, May 2022). Since the variant missingness rate was below 10% for all PRSs, we included all of them for testing. We calculated the individual scores in each of the five ancestries using the --score function in PLINK v1·9[13] and standardized them to mean zero and unit variance. Then, we tested the performance of the PRSs using the same procedure as for the validation cohorts.

To assess the added value of the PRS-CSx approach over constructing a PRS from meta-analyzed multi-ancestry GWAS results, we performed an IVW meta-analysis of D-PRISM ancestry-specific GWAS summary statistics and applied PRS-CS to the resulting meta-analysis. We used the EUR 1KG HM3 LD reference panel and allowed the shrinkage prior to be automatically learned from the data (which we refer to as 'D-PRISM multi-ancestry PRS-CS' model).

Similarly, we also evaluated the performance of a PRS based on the most statistically powered multi-ancestry T2D GWAS to date from Suzuki et al.[19,] including a 16% larger sample size than this study (i.e., Suzuki et al.: 2,535,601 individuals of which 428,452 are T2D cases and 2,107,149 controls vs. D-PRISM: 2,185,548 individuals of which 359,819 are T2D cases and 1,825,729 controls). We first applied the same quality control steps as those used for D-PRISM GWAS summary statistics and then used PRS-CS to construct a PRS using the EUR 1KG HM3 LD reference panel and let the prior shrinkage prior to be automatically learned from the data (which we refer to as 'Suzuki et al., PRS-CS' model).

Additionally, we constructed an rsPRS using the 1,289 distinct variants identified by Suzuki et al.,[19] defined as having an association $p<5 \times 10^{-8}$. For 90 out of 203 palindromic variants, we used proxy variants ($r^2 \geq 0·8$ in all the ancestry groups). We excluded 113 variants, as no proxy was available, thereby using 1,176 total variants to construct the rsPRS (which we refer to as 'Suzuki et al., rsPRS' model) (Supplementary Table 11).

Association of PRSs with diabetes complications

We also evaluated the association of the best-performing D-PRISM multi-ancestry PRS-CSx models with T2D-related microvascular complications (i.e., diabetic nephropathy, diabetic retinopathy, end-stage diabetic nephropathy, and proliferative diabetic retinopathy) and macrovascular complications (i.e., cardiovascular disease and ischemic stroke) in the AoU validation cohort. To comply with the AoU policies, we only considered individuals of the AFR, AMR, and EUR ancestry groups, as there were limited sample sizes for the EAS and SAS ancestries (i.e., <30 individuals with diabetes complications). We defined the traits based on ICD9 and ICD10 codes, as previously described.[19] We restricted microvascular complication analyses to individuals with T2D, as these outcomes are largely diabetes specific. For macrovascular complications, which also occur in those without T2D, we included all individuals and adjusted for T2D status in the models. We tested the association of each T2D-related complication with the standardized PRSs by fitting logistic regression models adjusted for sex, age, and genetics PCs in each ancestry group, separately (Supplementary Table 12).

Method's references

1 Mahajan A, Spracklen CN, Zhang W, *et al.* Multi-ancestry genetic study of type 2 diabetes highlights the power of diverse populations for discovery and translation. *Nat Genet* 2022; 54: 560–72.
2 Vujkovic M, Keaton JM, Lynch JA, *et al.* Discovery of 318 new risk loci for type 2 diabetes and related vascular outcomes among 1.4 million participants in a multi-ancestry meta-analysis. *Nat Genet* 2020; 52: 680–91.
3 Kurki MI, Karjalainen J, Palta P, *et al.* FinnGen provides genetic insights from a well-phenotyped isolated population. *Nature* 2023; 613: 508–18.
4 1000 Genomes Project Consortium, Auton A, Brooks LD, *et al.* A global reference for human genetic variation. *Nature* 2015; 526: 68–74.
5 Bergström A, McCarthy SA, Hui R, *et al.* Insights into human genetic variation and population history from 929 diverse genomes. *Science* 2020; 367. DOI:10.1126/science.aay5012.
6 Willer CJ, Li Y, Abecasis GR. METAL: fast and efficient meta-analysis of genomewide association scans. *Bioinformatics* 2010; 26: 2190–1.
7 Kowalski MH, Qian H, Hou Z, *et al.* Use of >100,000 NHLBI Trans-Omics for Precision Medicine (TOPMed) Consortium whole genome sequences improves imputation quality and detection of rare variant associations in admixed African and Hispanic/Latino populations. *PLoS Genet* 2019; 15: e1008500.
8 Taliun D, Harris DN, Kessler MD, *et al.* Sequencing of 53,831 diverse genomes from the NHLBI TOPMed Program. *Nature* 2021; 590: 290–9.
9 Das S, Forer L, Schönherr S, *et al.* Next-generation genotype imputation service and methods. *Nat Genet* 2016; 48: 1284–7.
10 Ge T, Chen C-Y, Ni Y, Feng Y-CA, Smoller JW. Polygenic prediction via Bayesian regression and continuous shrinkage priors. *Nat Commun* 2019; 10: 1776.
11 Ruan Y, Lin Y-F, Feng Y-CA, *et al.* Improving polygenic prediction in ancestrally diverse populations. *Nat Genet* 2022; 54: 573–80.
12 Berisa T, Pickrell JK. Approximately independent linkage disequilibrium blocks in human populations. *Bioinformatics* 2016; 32: 283–5.
13 Chang CC, Chow CC, Tellier LC, Vattikuti S, Purcell SM, Lee JJ. Second-generation PLINK: rising to the challenge of larger and richer datasets. *Gigascience* 2015; 4: 7.
14 Wojcik GL, Fuchsberger C, Taliun D, *et al.* Imputation-Aware Tag SNP Selection To Improve Power for Large-Scale, Multi-ethnic Association Studies. *G3 (Bethesda)* 2018; 8: 3255–67.
15 Robin X, Turck N, Hainard A, *et al.* pROC: an open-source package for R and S+ to analyze and compare ROC curves. *BMC Bioinformatics* 2011; 12: 77.
16 Viechtbauer W. Conducting Meta-Analyses in *R* with the metafor Package. *J Stat Softw* 2010; 36. DOI:10.18637/jss.v036.i03.
17 Lambert SA, Wingfield B, Gibson JT, *et al.* Enhancing the Polygenic Score Catalog with tools for score calculation and ancestry normalization. *Nat Genet* 2024; 56: 1989–94.
18 All of Us Research Program Genomics Investigators. Genomic data in the All of Us Research Program. *Nature* 2024; 627: 340–6.
19 Suzuki K, Hatzikotoulas K, Southam L, *et al.* Genetic drivers of heterogeneity in type 2 diabetes pathophysiology. *Nature* 2024; 627: 347–57.

### Supplementary Figures

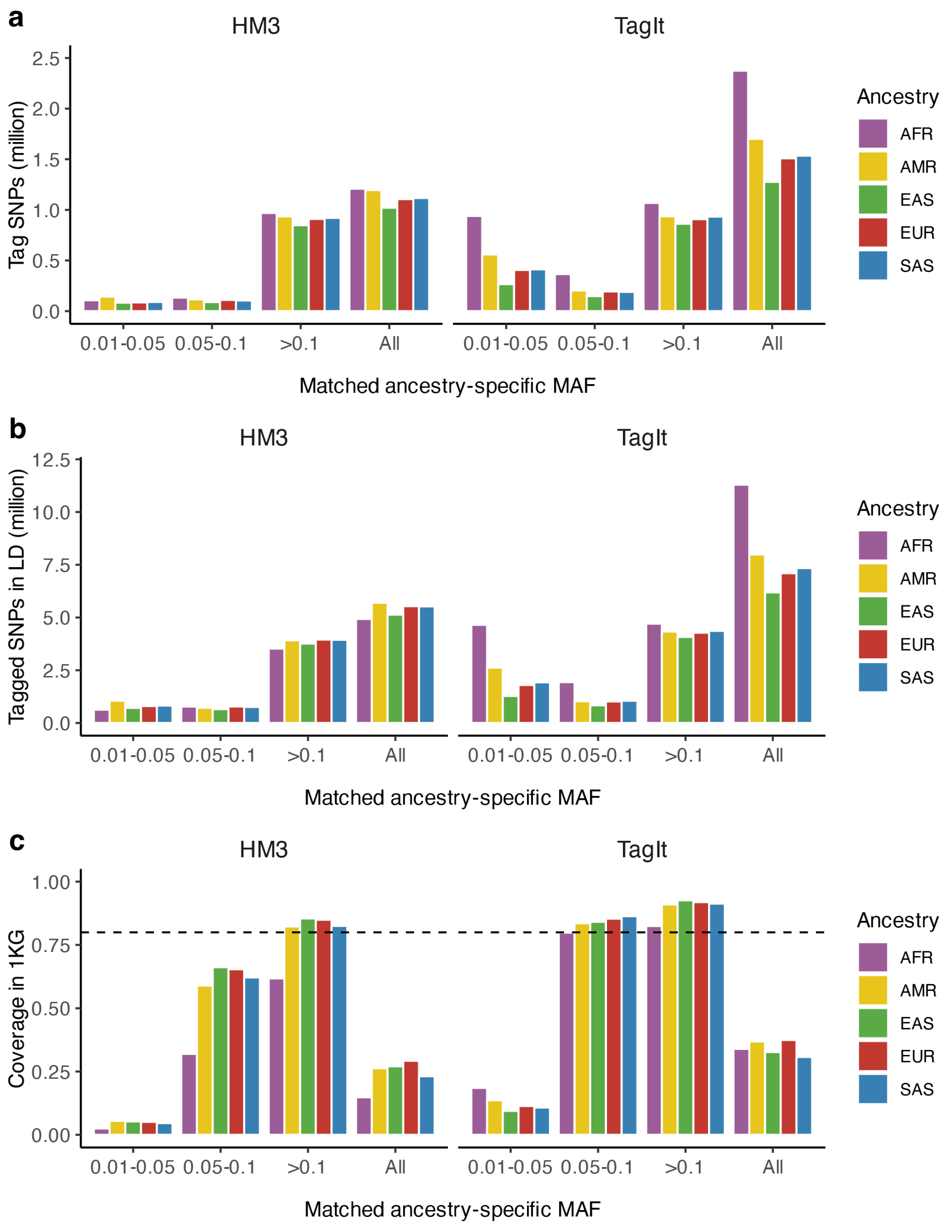

**Supplementary Fig.1 | Tag SNPs informativeness across ancestries.** We considered two sets of reference SNPs: one was based on the HapMap3 (HM3) set of variants, and the other was selected using the TagIt program. **a,** Number of tag SNPs in the LD reference panels, stratified by minor allele frequency and ancestry (*left*, HM3-based; *right*, TagIt-based), **b,** Number of SNPs being tagged in the LD reference panels, stratified by minor allele frequency and ancestry for a minimum pairwise correlation threshold of $r^2$>0.8 (*left*, HM3-based; *right*, TagIt-based), **c,** Proportion of SNPs that are either tags or are tagged ($r^2$>0.8) in the LD reference panels, stratified by minor allele frequency and ancestry in the individuals from 1KG. The dashed line represents 80% coverage.

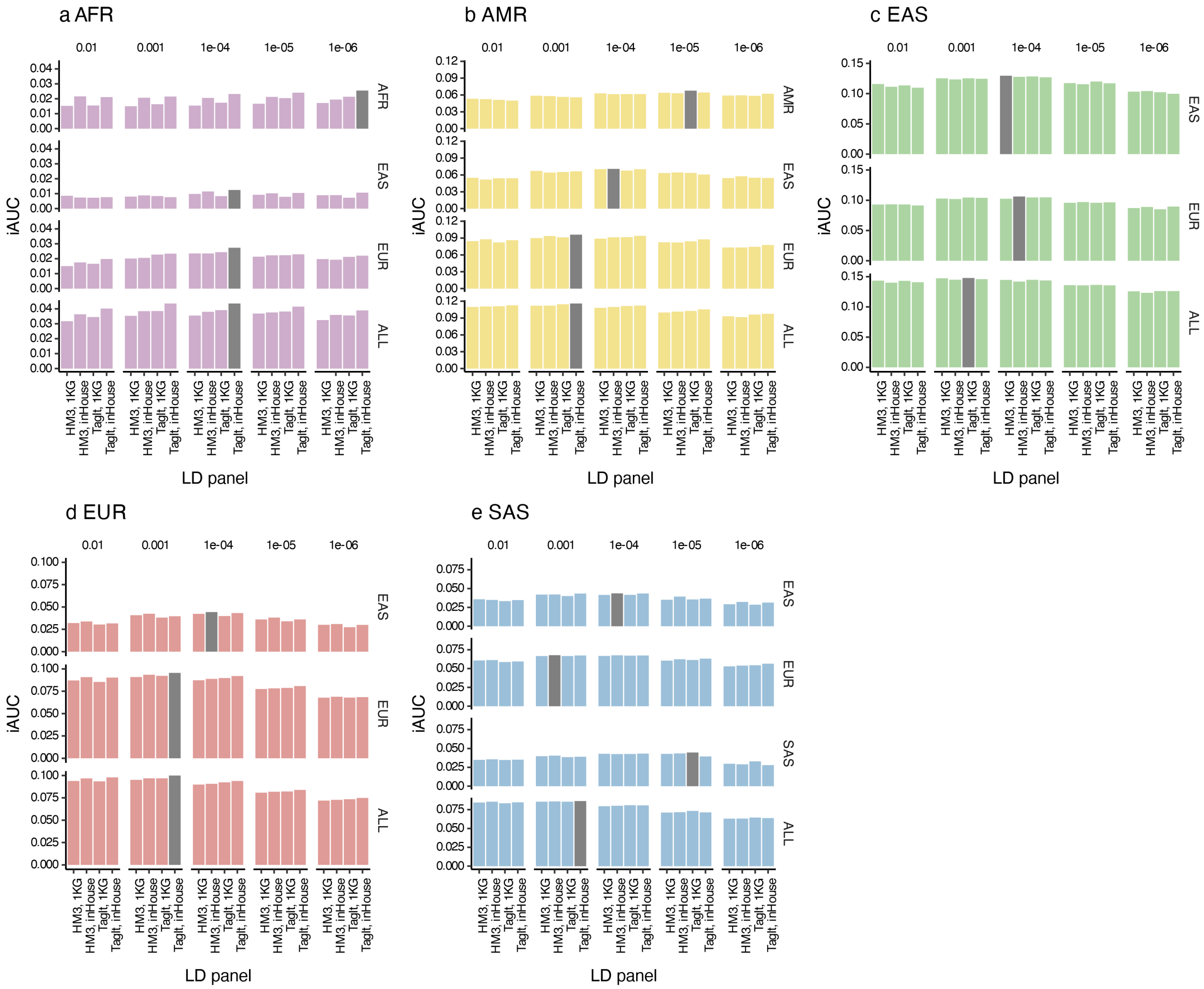

**Supplementary Fig.2 | Performance of the T2D PRSs in the training cohorts across ancestry groups.** Incremental AUC (iAUC) of the T2D PRS in the training cohorts: **a**, AFR, **b,** AMR, **c,** EAS, **d,** EUR, **e,** SAS. For each ancestry, we trained single-ancestry and multi-ancestry (ALL) PRSs using four LD panels and 5 phi continuous shrinkage priors. Bar colors represent the ancestry group: purple for AFR, yellow for AMR, green for EAS, red for EUR, and blue for SAS. The grey color highlights the best-trained PRS models that maximize the iAUC.

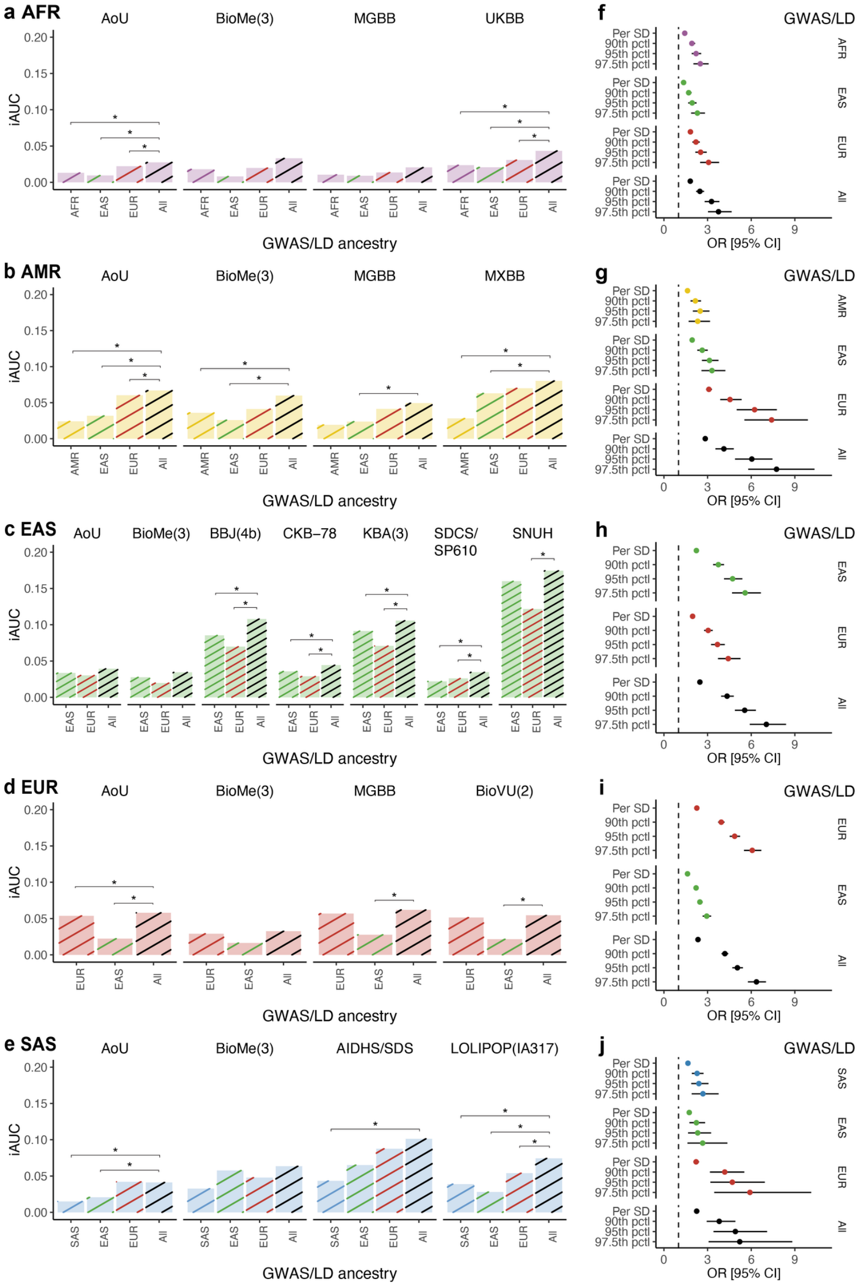

**Supplementary Fig. 3 | Performance of the T2D PRSs adjusted for BMI in the validation cohorts across ancestry groups. a-e:** Incremental AUC (iAUC) of the T2D PRS in the validation cohorts across ancestry groups: **a,** AFR, **b,** AMR, **c,** EAS, **d,** EUR, **e,** SAS. For each ancestry, the best-performing single-ancestry and multi-ancestry (All) PRSs were evaluated. Each bar represents a single cohort. Bar colors represent the ancestry group: purple for AFR, yellow for AMR, green for EAS, red for EUR, and blue for SAS. Line colors represent the ancestry of the T2D GWAS summary statistics and LD panels used to train the PRS, using the same color codes for single-ancestry PRSs, and black for multi-ancestry PRSs. **f-j:** Odds ratio (OR) from the meta-analysis of validation cohorts across ancestry groups: **f,** AFR, **g,** AMR, **h,** EAS, **i,** EUR, **j,** SAS. Points represent the odds ratio per standard deviation of the PRS distribution or the odds ratio comparing different PRS distribution extremes relative to the interquartile range. Error bars show the 95% confidence intervals (95% CI). Point colors represent the ancestry of the T2D GWAS summary statistics and LD panels used to train the PRS. * De Long $p<0.05$.

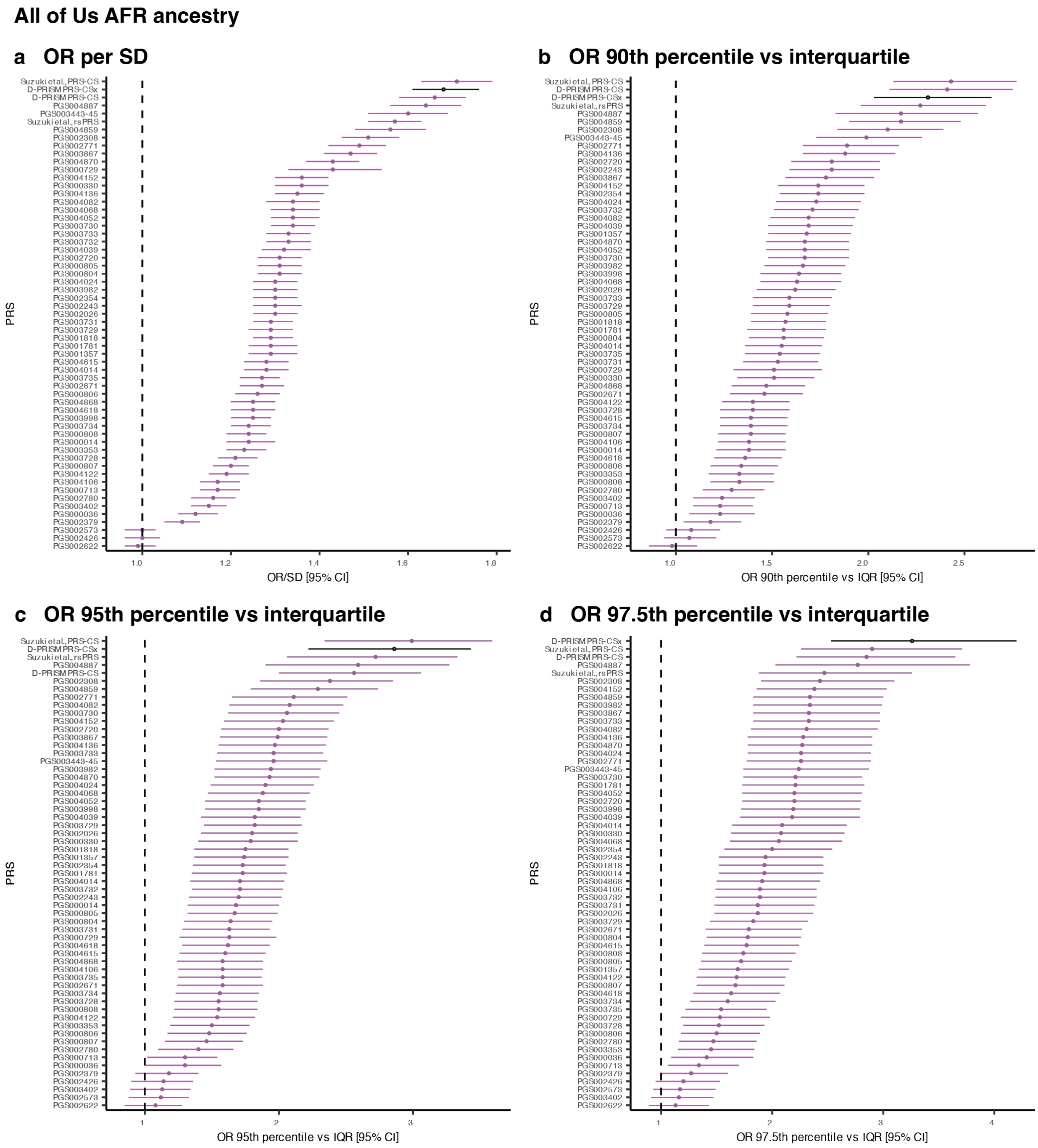

**Supplementary Fig.4 | Performance of D-PRISM multi-ancestry PRS-CSx compared to the published T2D PRSs from the PGS Catalog and others in individuals of AFR ancestry from the All of Us validation cohort. a,** Odds ratio per standard deviation (OR per SD) of the PRS distribution, **b,** OR comparing the 90th percentile of the PRS relative to the interquartile range, **c,** OR comparing the 95th percentile of the PRS relative to the interquartile range, **d,** OR comparing the 97.5th percentile of the PRS relative to the interquartile range.

### All of Us AMR ancestry

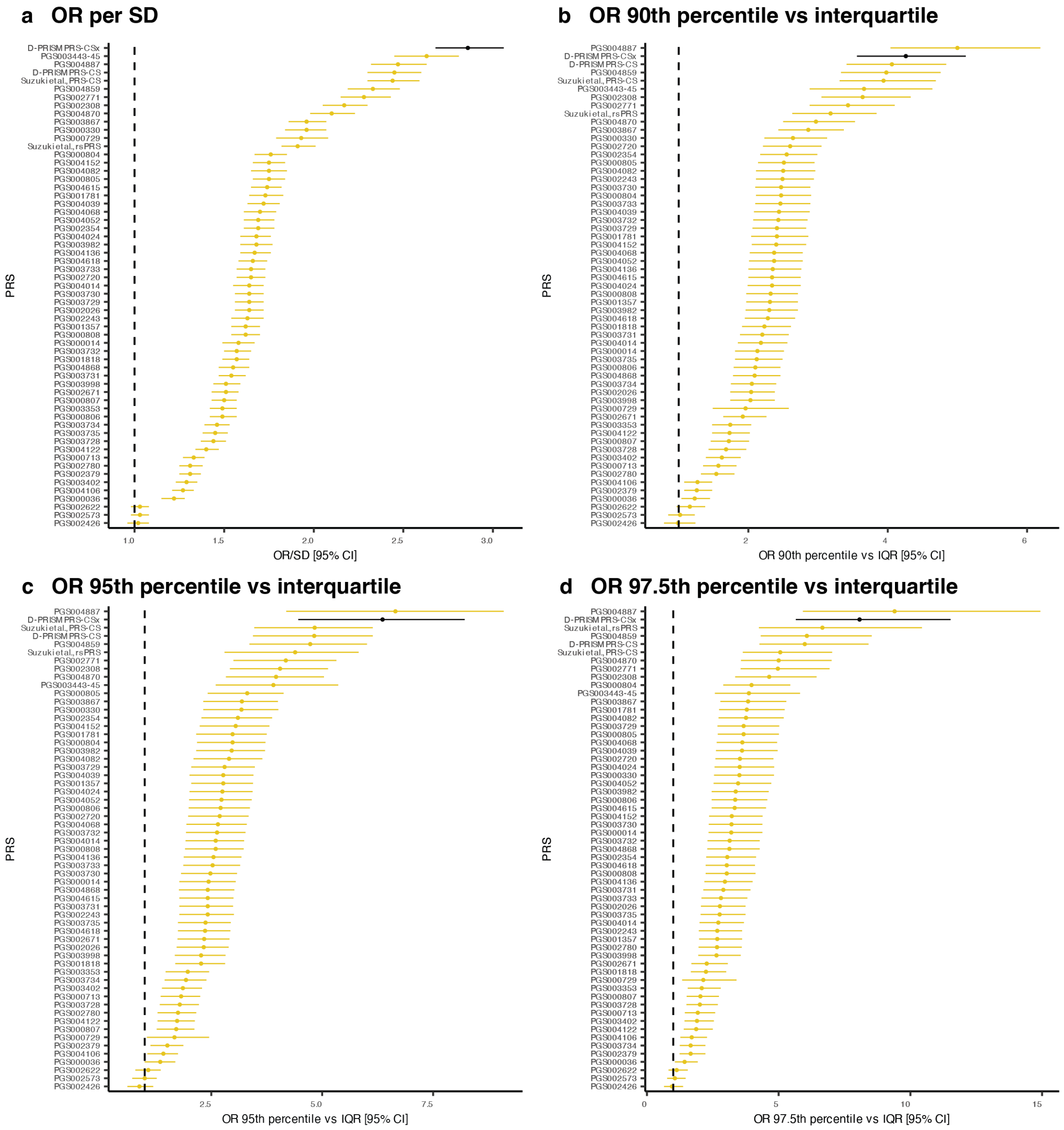

**Supplementary Fig.5 | Performance of D-PRISM multi-ancestry PRS-CSx compared to the published T2D PRSs from the PGS Catalog and others in individuals of AMR ancestry from the All of Us validation cohort. a,** Odds ratio per standard deviation (OR per SD) of the PRS distribution, **b,** OR comparing the 90$^{th}$ percentile of the PRS relative to the interquartile range, **c,** OR comparing the 95$^{th}$ percentile of the PRS relative to the interquartile range, **d,** OR comparing the 97.5$^{th}$ percentile of the PRS relative to the interquartile range.

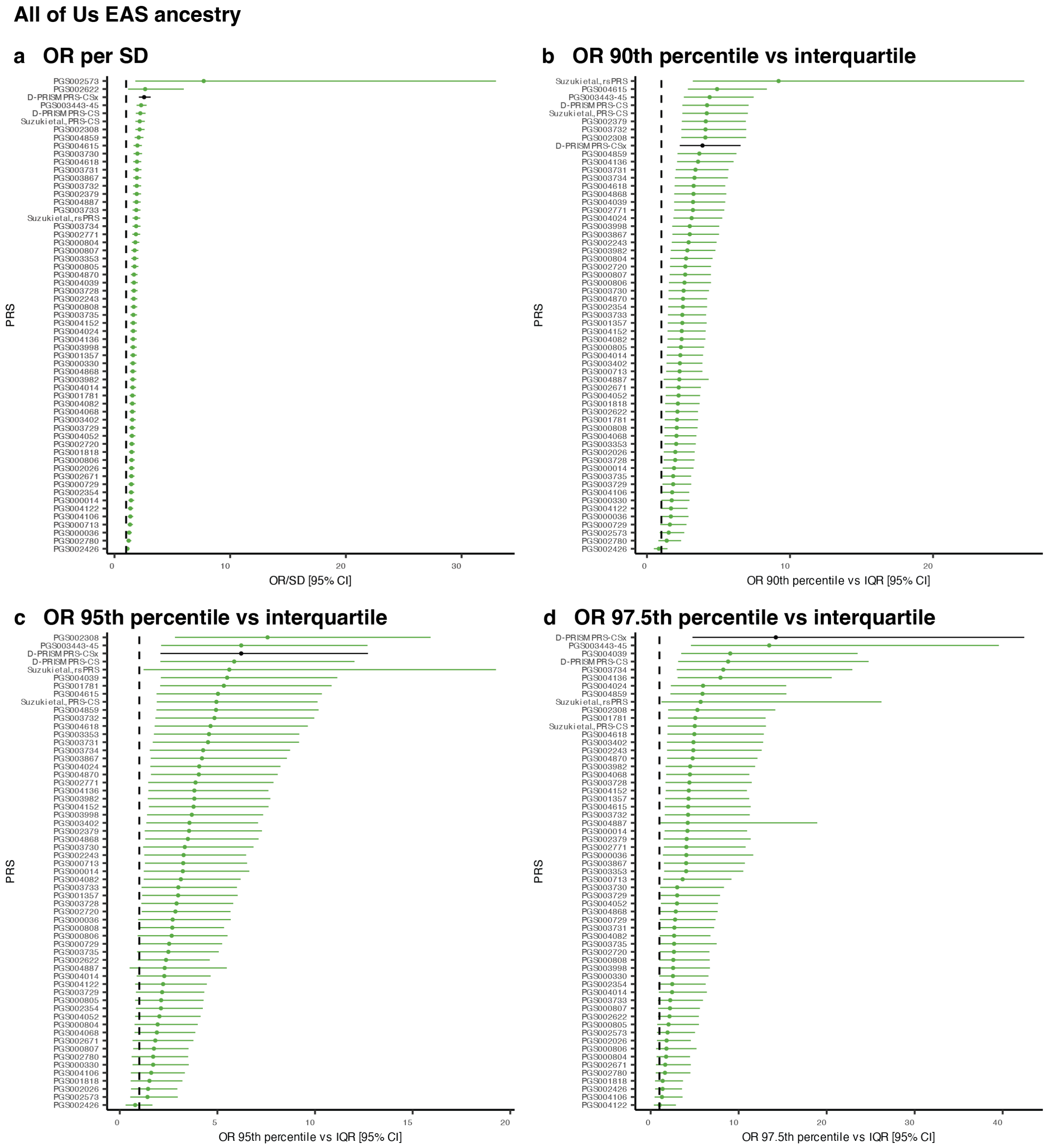

**Supplementary Fig.6 | Performance of D-PRISM multi-ancestry PRS-CSx compared to the published T2D PRSs from the PGS Catalog and others in individuals of EAS ancestry from the All of Us validation cohort. a,** Odds ratio per standard deviation (OR per SD) of the PRS distribution, **b,** OR comparing the 90$^{th}$ percentile of the PRS relative to the interquartile range, **c,** OR comparing the 95$^{th}$ percentile of the PRS relative to the interquartile range, **d,** OR comparing the 97.5$^{th}$ percentile of the PRS relative to the interquartile range.

### All of Us EUR ancestry

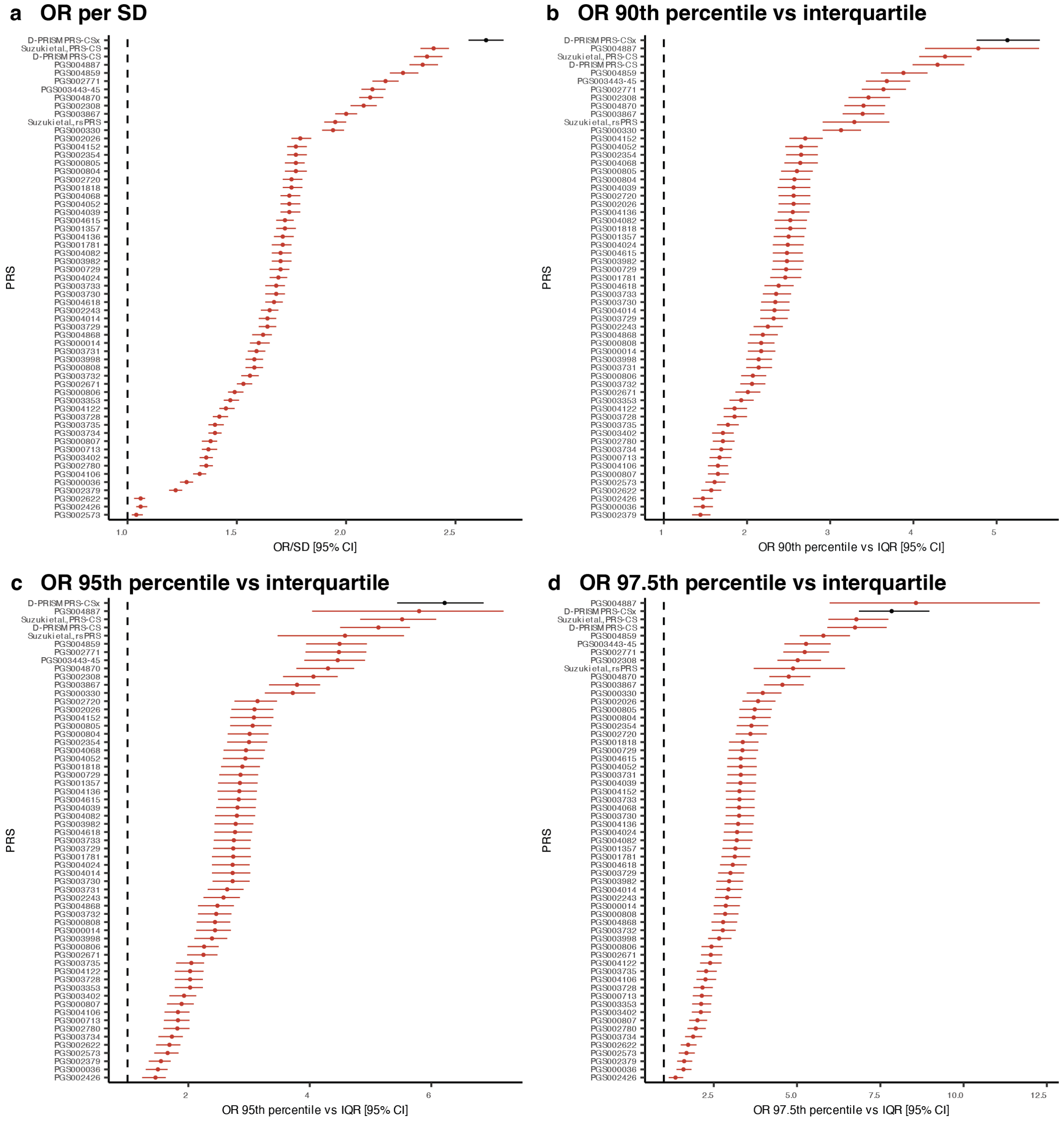

**Supplementary Fig.7 | Performance of D-PRISM multi-ancestry PRS-CSx compared to the published T2D PRSs from the PGS Catalog and others in individuals of EUR ancestry from the All of Us validation cohort. a,** Odds ratio per standard deviation (OR per SD) of the PRS distribution, **b,** OR comparing the 90th percentile of the PRS relative to the interquartile range, **c,** OR comparing the 95th percentile of the PRS relative to the interquartile range, **d,** OR comparing the 97.5th percentile of the PRS relative to the interquartile range.

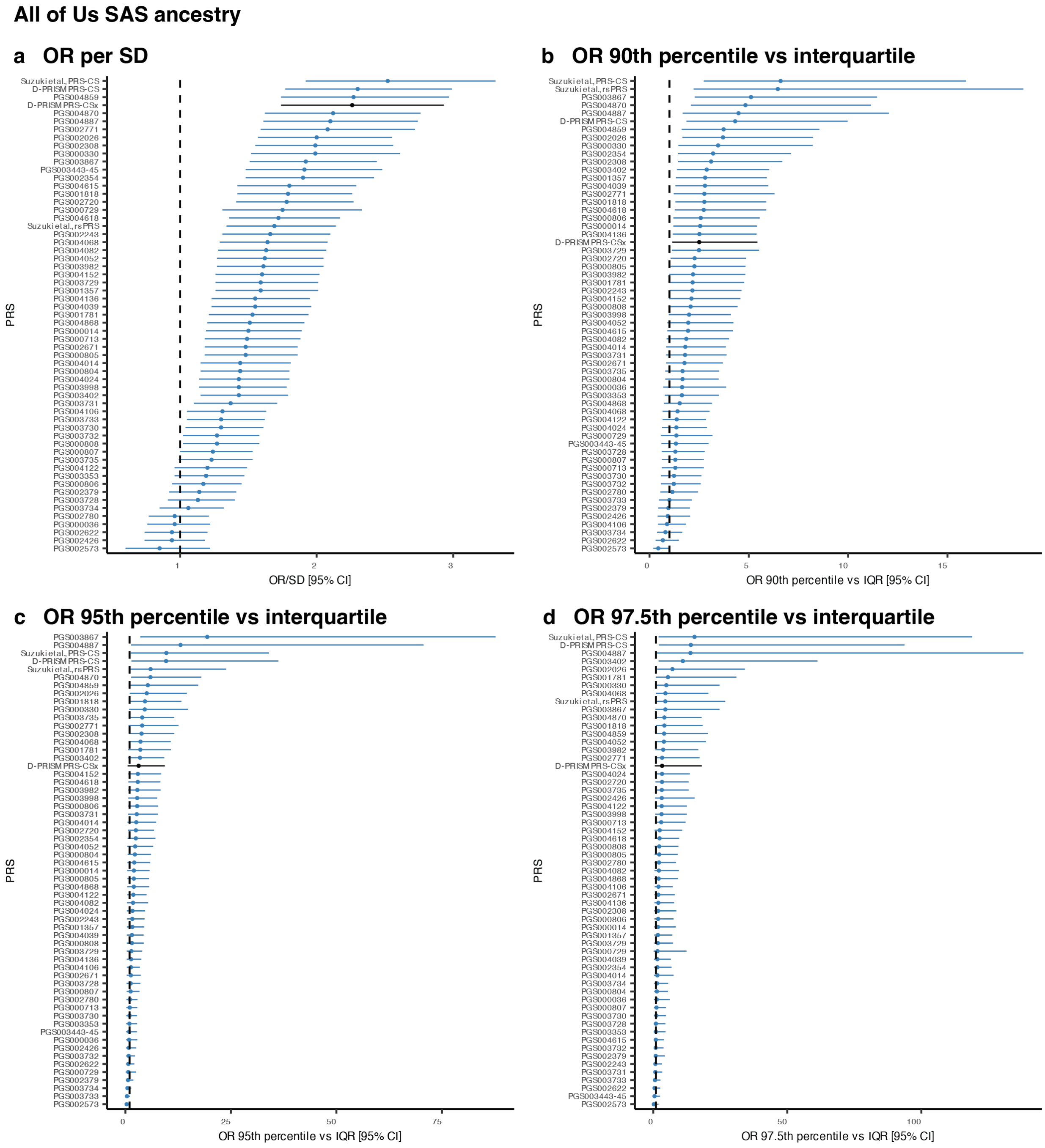

**Supplementary Fig.8 | Performance of D-PRISM multi-ancestry PRS-CSx compared to the published T2D PRSs from the PGS Catalog and others in individuals of SAS ancestry from the All of Us validation cohort. a,** Odds ratio per standard deviation (OR per SD) of the PRS distribution, **b,** OR comparing the 90$^{th}$ percentile of the PRS relative to the interquartile range, **c,** OR comparing the 95$^{th}$ percentile of the PRS relative to the interquartile range, **d,** OR comparing the 97.5$^{th}$ percentile of the PRS relative to the interquartile range.

**Cohort acknowledgments and funding**

Anti-aging Study Cohort (AASC) is supported by the Grant-in-Aid for Scientific Research (20018020, 19659163, 20390185, 23659382, 24390084, 23659352, 25293141, 26670313, 17H04123) from the Ministry of Education, Culture, Sports, Science and Technology of Japan, research grant from the Japan Atherosclerosis Prevention Found, National Cardiovascular Research Grants, and Research Promotion Award from Ehime University.

Asian Indian Diabetic Heart Study/Sikh Diabetes Study (AIDHS/SDS) was started in 2002 by the Fogarty International Center of the National Institute of Health (NIH), NIH/FIC K01 TW006087 (2002-2005) and 2K01 TW006087 (2006-2010) awards. The AIDHS/SDS has been supported by NIH Grants: NHLBI -HV48141, NHLBI R&G Service/R218, NIDDK/R01DK082766, NIH/NHGRI HG-11-009, NIDDK/DK105913, NIDDK/R01DK118427. The authors thank the participants of the study for their important contributions.

All Of Us Research Program (AOU) is supported by the National Institutes of Health, Office of the Director: Regional Medical Centers: 1 OT2 OD026549; 1 OT2 OD026554; 1 OT2 OD026557; 1 OT2 OD026556; 1 OT2 OD026550; 1 OT2 OD 026552; 1 OT2 OD026553; 1 OT2 OD026548; 1 OT2 OD026551; 1 OT2 OD026555; IAA #: AOD 16037; Federally Qualified Health Centers: HHSN 263201600085U; Data and Research Center: 5 U2C OD023196; Biobank: 1 U24 OD023121; The Participant Center: U24 OD023176; Participant Technology Systems Center: 1 U24 OD023163; Communications and Engagement: 3 OT2 OD023205; 3 OT2 OD023206; and Community Partners: 1 OT2 OD025277; 3 OT2 OD025315; 1 OT2 OD025337; 1 OT2 OD025276. In addition, the All of Us Research Program would not be possible without the partnership of its participants.

Atherosclerosis Risk in Communities (ARIC) study has been funded in whole or in part with Federal funds from the National Heart, Lung, and Blood Institute, National Institutes of Health, Department of Health and Human Services (contract numbers HHSN268201700001I, HHSN268201700002I, HHSN268201700003I, HHSN268201700004I and HHSN268201700005I), R01HL087641, R01HL059367 and R01HL086694; National Human Genome Research Institute contract U01HG004402; and National Institutes of Health contract HHSN268200625226C. The authors thank the staff and participants of the ARIC study for their important contributions. Infrastructure was partly supported by Grant Number UL1RR025005, a component of the National Institutes of Health and NIH Roadmap for Medical Research.

BioBank Japan (BBJ). This study was funded by the BioBank Japan project, which is supported by the Ministry of Education, Culture, Sports, Sciences and Technology (MEXT) of Japanese government and the Japan Agency for Medical Research and Development (AMED, grant ID JP21km0605001). AMED GRIFIN Diabetes Initiative Japan was supported by Japan Agency for Medical Research and Development (JP20km0405202, JP21tm0424218). Scarda was supported by AMED under Grant Number 223fa627011.

Beijing Eye Study (BES) was supported by National Natural Science Foundation of China (grant 81570835).

BioMe Biobank (BIOME) is supported by The Andrea and Charles Bronfman Philanthropies and in part by funding of the NIH (U01HG007417; R56HG010297; X01HL134588). BIOME thanks all participants in the Mount Sinai Biobank, and also thanks all the recruiters who have assisted and continue to assist in data collection and management. BIOME is grateful for the computational resources and staff expertise provided by Scientific Computing at the Icahn School of Medicine at Mount Sinai.

Vanderbilt University Medical Center (BIOVU) projects are supported by numerous sources: institutional funding, private agencies, and federal grants. These include NIH funded Shared Instrumentation Grant S10OD017985, S10RR025141, and S10OD025092; CTSA grants UL1TR002243, UL1TR000445, and UL1RR024975. Genomic data are also supported by investigator-led projects that include U01HG004798, R01NS032830, RC2GM092618, P50GM115305, U01HG006378, U19HL065962, and R01HD074711. This work was conducted in part using the resources of the Advanced Computing Center for Research and Education at Vanderbilt University, Nashville, TN, supported in part by an S10 instrumentation award (1S10OD023680-01).

Bangladesh Population Cohort (BPC) was supported by US National Institute of Environmental Health Sciences Grants P42 ES10349 and P30 ES09089. Cardiometabolic Genome Epidemiology (CAGE-AMAGASKI and CAGE-

GWAS) was supported by grants for the Core Research for Evolutional Science and Technology (CREST) from the Japan Science Technology Agency; KAKENHI (Grant-in-Aid for Scientific Research) from the Ministry of Education, Culture, Sports, Science and Technology of Japan; and the Grant and research budget of National Center for Global Health and Medicine (NCGM). CAGE-AMAGASKI thanks Drs. Toshio Ogihara, Yukio Yamori, Akihiro Fujioka, Chikanori Makibayashi, Sekiharu Katsuya, Ken Sugimoto, Kei Kamide, and Ryuichi Morishita and the many physicians of the participating hospitals and medical institutions in Amagasaki Medical Association for their assistance in collecting the DNA samples and accompanying clinical information.

Cardiometabolic Genome Epidemiology Kita-Nagoya Genomic Epidemiology (CAGE-KING) was supported in part by Grants-in-Aid from MEXT (nos. 24390169, 16H05250, 15K19242, 16H06277) as well as by a grant from the Funding Program for Next-Generation World-Leading Researchers (NEXT Program, no. LS056).

Coronary Artery Risk Development in Young Adults (CARDIA) was conducted and supported by the National Heart, Lung, and Blood Institute (NHLBI) in collaboration with the University of Alabama at Birmingham (HHSN268201800005I & HHSN268201800007I), Northwestern University (HHSN268201800003I), University of Minnesota (HHSN268201800006I), and Kaiser Foundation Research Institute (HHSN268201800004I). CARDIA was also partially supported by the Intramural Research Program of the National Institute on Aging (NIA) and an intra-agency agreement between NIA and NHLBI (AG0005). Genotyping was funded as part of the NHLBI Candidate-gene Association Resource (N01-HC-65226) and the NHGRI Gene Environment Association Studies (GENEVA) (U01-HG004729, U01-HG04424, and U01-HG004446).

Cleveland Family Study (CFS) is supported by grants to Case Western Reserve University (NIH HL 46380, M01RR00080) and Brigham and Women's Hospital (K01-HL135405-01, R01-HL113338-04, R35-HL135818-01, 5-R01-HL046380-15 and 5-KL2-RR024990-05).

China Health and Nutrition Survey (CHNS) was supported by: the National Institute for Nutrition and Health, the Chinese Center for Disease Control and Prevention; the National Institutes of Health (R01AG065357, R01HD30880, R01HL108427 and R01DK104371); the Fogarty International Center of the National Institutes of Health (TW009077); the China-Japan Friendship Hospital, the Beijing Municipal Center for Disease Prevention and Control, the China National Health Commission (formerly the Chinese Ministry of Health); the Chinese National Human Genome Center at Shanghai; and the Carolina Population Center (P2CHD050924), The University of North Carolina at Chapel Hill.

Cardiovascular Health Study (CHS). This Cardiovascular Health Study research was supported by NHLBI contracts HHSN268201200036C, HHSN268200800007C, HHSN268201800001C, N01HC55222, N01HC85079, N01HC85080, N01HC85081, N01HC85082, N01HC85083, N01HC85086, 75N92021D00006; and NHLBI grants U01HL080295, R01HL087652, R01HL103612, R01HL105756, R01HL120393, U01HL130114, and R01HL172803 with additional contribution from the National Institute of Neurological Disorders and Stroke (NINDS). Additional support was provided through R01AG023629 from the National Institute on Aging (NIA). A full list of principal CHS investigators and institutions can be found at CHS-NHLBI.org. The provision of genotyping data was supported in part by the National Center for Advancing Translational Sciences, CTSI grant UL1TR001881, and the National Institute of Diabetes and Digestive and Kidney Disease Diabetes Research Center (DRC) grant DK063491 to the Southern California Diabetes Endocrinology Research Center.

China Kadoorie Biobank (CKB) chiefly acknowledges the participants, project staff, and the China National Centre for Disease Control and Prevention (CDC) and its regional offices. China's National Health Insurance provides electronic linkage to all hospital treatment. Funding sources: Baseline survey and first re-survey - Kadoorie Charitable Foundation, Hong Kong; long-term follow-up - UK Wellcome Trust (212946/Z/18/Z, 202922/Z/16/Z, 104085/Z/14/Z, 088158/Z/09/Z), National Natural Science Foundation of China (82192901, 82192904, 82192900), and National Key Research and Development Program of China (2016YFC 0900500, 0900501, 0900504, 1303904); DNA extraction and genotyping –GlaxoSmithKline, and the UK Medical Research Council (MC-PC-13049, MC-PC-14135); core funding for the project to the Clinical Trial Service Unit and Epidemiological Studies Unit at Oxford University - British Heart Foundation (CH/1996001/9454), UK MRC (MC-UU-00017/1, MC-UU-12026/2, MC_U137686851), and Cancer Research UK (C16077/A29186, C500/A16896).

Cebu Longitudinal Health and Nutrition Survey (CLHNS) was supported by: US National Institutes of Health grants DK078150, TW005596 and HL085144; pilot funds from RR020649, ES010126, and DK056350; and the Office of Population Studies Foundation.

Diabetic Cohort and Singapore Prospective Study Program (DC/SP2) were supported by the individual research grant and clinician scientist award schemes from the National Medical Research Council (NMRC) and the Biomedical Research Council (BMRC) of Singapore, Ministry of Health, Singapore, and infrastructure funding from the Singapore Ministry of Health (Population Health Metrics and Analytics PHMA), National University of Singapore and National University Health System, Singapore.

Durban Diabetes Study and Durban Diabetes Case Control (DDS/DCC) was supported by: the Wellcome Trust (grant number 098051); the African Partnership for Chronic DiseaseResearch (Medical Research Council UK partnership grant number MR/K013491/1); the National Institute for Health Research Cambridge Biomedical Research Centre (UK); Novo-Nordisk (South Africa); Sanofi-Aventis (South Africa); MSD Pharmaceuticals (Pty) Ltd (Southern Africa); Servier Laboratories (South Africa); South African Sugar Association; and the Victor Daitz Foundation.

deCODE genetics (DECODE) thank the participants in the deCODE study, the staff at deCODE genetics core facilities and the staff at the Research Service Center for their contribution to this work.

Diabetes Gene Discovery Group (DGDG) was supported by Genome Canada, Génome Québec, the Canada Foundation for Innovation, the French Government (“Agence Nationale de la Recherche”), the French Region of "Nord Pas De Calais" ("Contrat de Projets État-Région"), and the charities: “Association Française des Diabétiques”, “Programme National de Recherche sur le Diabète” and “Association de Langue Française pour l'Etude du Diabète et des Maladies Métaboliques”. This study was also supported in part by a grant from the European Union (Integrated Project EuroDia LSHM-CT-2006-518153in the Framework Programme 6 [FP6] of the European Community). This work was supported by grants from the French National Research Agency (ANR-10-LABX-46 [European Genomics Institute for Diabetes] and ANR-10-EQPX-07-01 [LIGAN-PM]). Case and control recruitment was supported by the Fédération Française des Diabetiques, INSERM, CNAMTS, Centre Hospitalier Universitaire Poitiers, La Fondation de France, and the Endocrinology-Diabetology department of the Corbeil-Essonnes Hospital. C. Petit, J.-P. Riveline, and S. Franc were instrumental in recruitment and S. Brunet, F. Bacot, R. Frechette, V. Catudal, M. Deweirder, F. Allegaert, P. Laflamme, P. Lepage, W. Astle, M. Leboeuf, and S. Leroux provided technical assistance. K. Shazand and N. Foisset provided organizational guidance. The D.E.S.I.R. study, which mostly contributed controls, was supported by CNAMTS, Lilly, Novartis Pharma and Sanofi-Aventis, by INSERM (“Réseaux en Santé Publique, Interactions entre les déterminants de la santé”), by “Association Diabète Risque Vasculaire”, “Fédération Française de Cardiologie”, “Fondation de France”, ALFEDIAM, ONIVINS, Ardix Medical, Bayer Diagnostics, Becton Dickinson, Cardionics, Merck Santé, Novo Nordisk, Pierre Fabre, Roche, Topcon. The D.E.S.I.R. Study Group: INSERM U780: B. Balkau, P. Ducimetière, E. Eschwège; INSERM U367: F. Alhenc-Gelas; CHU D'Angers: Y. Gallois, A. Girault; Bichat Hospital: F. Fumeron, M. Marre; Medical Examination Services: Alençon, Angers, Caen, Chateauroux, Cholet, Le Mans, and Tours; Research Institute for General Medicine: J. Cogneau; General practitioners of the region; Cross-Regional Institute for Health: C. Born, E. Caces, M. Cailleau, J. G. Moreau, F. Rakotozafy, J. Tichet, S. Vol. DGDG thank M. Deweider and F. Allegaert for the DNA bank management and are sincerely indebted to all study participants.

Diabetes Genetics Initiative (DGI) was supported by the Novartis Institute for BioMedical Research with additional support from The Richard and Susan Smith Family Foundation and American Diabetes Association Pinnacle Program Project Award. The Botnia Study (study subject cohort) was financially supported by the Folkhalsan Research Foundation, the Sigrid Juselius Foundation, Nordic Center of Excellence in Disease Genetics, EU (EXGENESIS), The Academy of Finland, University of Helsinki, Finnish Diabetes Research Foundation, Foundation for Life and Health in Finland, Finnish Medical Society, Helsinki UniversityCentral Hospital Research Foundation, Perklén Foundation, Ollqvist Foundation, Närpes Health Care Foundation, Municipal Heath Care Center and Hospital in Jakobstad and Health Care Centers in Vasa, Närpes and Korsholm. The work in Malmö, Sweden, was also funded by a Linné grant from the Swedish Research Council (349-2006-237). The contribution of the Botnia and Skara research teams is gratefully acknowledged.

Electronic Medical Records and Genomics Network (EMERGE) was initiated and funded by NHGRI through the following grants: U01HG006828 (Cincinnati Children’s Hospital Medical Center/Boston Children’s Hospital);

U01HG006830 (Children’s Hospital of Philadelphia); U01HG006389 (Essentia Institute of Rural Health, Marshfield Clinic Research Foundation and Pennsylvania State University); U01HG006382 (Geisinger Clinic); U01HG006375 (Group Health Cooperative/University of Washington); U01HG006379 (Mayo Clinic); U01HG006380 (Icahn School of Medicine at Mount Sinai); U01HG006388 (Northwestern University); U01HG006378 (Vanderbilt University Medical Center); and U01HG006385 (Vanderbilt University Medical Center serving as the Coordinating Center). The Northwestern University Enterprise Data Warehouse was funded in part by a grant from the National Center for Research Resources, UL1RR025741. Part of the dataset(s) used for the analyses described were obtained from Vanderbilt University Medical Center's BioVU which is supported by institutional funding and by the Vanderbilt CTSA grant UL1 TR000445 from NCATS/NIH. The eMERGE imputed merged Phase I and Phase II dataset was generated by genotyping centers CIDR (U01HG004438) and the Broad Institute (U01HG004424).

European Prospective Investigation into Cancer and Nutrition (EPIC-INTERACT) project (LSHM-CT-2006-037197) is a European-Community funded project under Framework Programme 6. EPIC-INTERACT thank all EPIC participants and staff for their contribution to the study. EPIC-INTERACT thank Nicola Kerrison (MRC Epidemiology Unit, Cambridge) for managing the data for the InterAct Project and staff from the Laboratory Team, Field Epidemiology Team, and Data Functional Group of the MRC Epidemiology Unit in Cambridge, UK, for carrying out sample preparation, DNA provision and quality control, genotyping, and data-handling work. The funders had no role in study design, data collection and analysis, decision to publish, or preparation of the manuscript. GWAS summary statistics from the EPIC-InterAct study are available to download from the Dryad Digital Repository (https://doi.org/10.5061/dryad.qnk98sfcg).

Epidemiologic Study of the Screenees for Diabetes Reduction Assessment with Ramipril and Rosiglitazone Medication (EPIDREAM) was funded by a grant from the Canadian Institutes of Health Research University Industry competition with partner funding from the GlaxoSmithKline and Sanofi Aventis Global, Sanofi Aventis Canada, Genome Quebec Innovation Centre, Heart and Stroke Foundation of Canada.

Estonian Biobank (ESTBB) was funded by the Estonian Research Council Grant IUT20-60, IUT24-6, PRG687, and the European Union through the European Regional Development Fund Project No. 2014-2020.4.01.15-0012 GENTRANSMED.

Family Heart Study (FAMHS) was supported by NIH grants R01-HL-087700 and R01-HL-088215 from NHLBI, and R01-DK-089256 and R01-DK-075681 from NIDDK.

Framingham Heart Study (FHS) was conducted and supported by the National Heart, Lung and Blood Institute (NHLBI) in collaboration with Boston University (contracts 75N92019D00031, HHSN268201500001I and N01-HC-25195), and its contract with Affymetrix, Inc for genotyping services (contract number N02-HL-6-4278). The analyses reflect intellectual input and resource development from the Framingham Heart Study investigators participating in the SNP Health Association Resource (SHARe) project. FHS was also supported by: NHLBI R01 HL105756, National Institute for Diabetes and Digestive and Kidney Diseases (NIDDK) R01 DK078616, U01 DK078616, NIDDK K24 DK080140 and American Diabetes Association Mentor-Based Postdoctoral Fellowship Award #7-09-MN-32 (to J.B.M.); and NIDDK K24 DK110550 (to J.C.F.).

FinnGen study (FINNGEN) is a large-scale genomics initiative that has analyzed over 500,000 Finnish biobank samples and correlated genetic variation with health data to understand disease mechanisms and predispositions. The project is a collaboration between research organisations and biobanks within Finland and international industry partners. We want to acknowledge the participants and investigators of the FinnGen study.

Finland-United States Investigation of NIDDM Genetics (FUSION) was supported by DK093757, DK072193, DK062370, and ZIA-HG000024.

German Chronic Kidney Disease (GCKD) was funded by the German Ministry of Research and Education (Bundesminsterium für Bildung und Forschung, BMBF) and by the Foundation KfH Stiftung Präventivmedizin. Unregistered grants to support the study were provided by Bayer, Fresenius Medical Care and Amgen. Genotyping was supported by Bayer AG.

Genetic Study of Atherosclerosis Risk (GENESTAR) was supported by NIH grants through the National Heart, Lung, and Blood Institute (HL49762, HL58625, HL59684, HL071025, U01HL72518, and HL087698) and the National Institute of Nursing Research (NR0224103) and by M01-RR000052 to the Johns Hopkins General Clinical Research Center. Genetic Epidemiology Network of Arteriosclerosis (GENOA) was supported by the National Institutes of Health grant numbers HL054457, HL054464, HL054481, HL087660 and HL119443 from the National Heart, Lung, and Blood Institute. Genotyping was performed at the Mayo Clinic by Stephen Turner, Mariza de Andrade, and Julie Cunningham. GENOA thanks Eric Boerwinkle and Megan Grove from the Human Genetics Center and Institute of Molecular Medicine and Division of Epidemiology, University of Texas Health Science Center, Houston, Texas, USA for their help with genotyping. GENOA also thanks the families that participated in the study.

Genes and Health. Genes & Health is/has recently been core-funded by Wellcome (WT102627, WT210561), the Medical Research Council (UK) (M009017, MR/X009777/1, MR/X009920/1), Higher Education Funding Council for England Catalyst, Barts Charity (845/1796), Health Data Research UK (for London substantive site), and research delivery support from the NHS National Institute for Health Research Clinical Research Network (North Thames). Genes & Health is/has recently been funded by Alnylam Pharmaceuticals, Genomics PLC; and a Life Sciences Industry Consortium of AstraZeneca PLC, Bristol-Myers Squibb Company, GlaxoSmithKline Research and Development Limited, Maze Therapeutics Inc, Merck Sharp & Dohme LLC, Novo Nordisk A/S, Pfizer Inc, Takeda Development Centre Americas Inc. We thank Social Action for Health, Centre of The Cell, members of our Community Advisory Group, and staff who have recruited and collected data from volunteers. We thank the NIHR National Biosample Centre (UK Biocentre), the Social Genetic & Developmental Psychiatry Centre (King's College London), Wellcome Sanger Institute, and Broad Institute for sample processing, genotyping, sequencing and variant annotation. This work uses data provided by patients and collected by the NHS as part of their care and support. This research utilised Queen Mary University of London's Apocrita HPC facility, supported by QMUL Research-IT, http://doi.org/10.5281/zenodo.438045. We thank: Barts Health NHS Trust, NHS Clinical Commissioning Groups (City and Hackney, Waltham Forest, Tower Hamlets, Newham, Redbridge, Havering, Barking and Dagenham), East London NHS Foundation Trust, Bradford Teaching Hospitals NHS Foundation Trust, Public Health England (especially David Wyllie), Discovery Data Service/Endeavour Health Charitable Trust (especially David Stables), Voror Health Technologies Ltd (especially Sophie Don), NHS England (for what was NHS Digital) - for GDPR-compliant data sharing backed by individual written informed consent. Most of all we thank all of the volunteers participating in Genes & Health. A favourable ethical opinion for the main Genes & Health research study was granted by NRES Committee London - South East (reference 14/LO/1240) on 16 Sept 2014. Queen Mary University of London is the Sponsor.

Resource for Genetic Epidemiology on Adult Heath and Aging (GERA) was supported by a grant (RC2 AG033067; PIs Schaefer and Risch) awarded to the Kaiser Permanente Research Program on Genes, Environment, and Health (RPGEH) and the UCSF Institute for Human Genetics. The RPGEH was supported by grants from the Robert Wood Johnson Foundation, the Wayne and Gladys Valley Foundation, the Ellison Medical Foundation, Kaiser Permanente Northern California, and the Kaiser Permanente National and Northern California Community Benefit Programs.

Genetics of Diabetes and Audit Research in Tayside Scotland (GODARTS) was funded by The Wellcome Trust Study Cohort Functional Genomics Grant (2004-2008, 072960/Z/03/Z) and The Wellcome Trust Scottish Health Informatics Programme (SHIP, 2009-2012, 086113/Z/08/Z).

Genetics of Latinos Diabetic Retinopathy (GOLDR) was supported by grants EY14684 and UL1TR000124.

Genetic Overlap Between Metabolic and Psychiatric Traits and Teens of Attica: Genes and Environment (GOMAP-TEENAGE) was funded by the Wellcome Trust (098051) and was also co-financed by the European Union (European Social Fund - ESF) and Greek national funds through the Operational Program "Education and Lifelong Learning" of the National Strategic Reference Framework (NSRF) - Research Funding Program: Heracleitus II. GOMAP-TEENAGE thanks all study participants and their families, as well as all volunteers for their contribution in this study. GOMAP-TEENAGE is grateful to: Georgia Markou, Laiko General Hospital Diabetes Centre; Maria Emetsidou and Panagiota Fotinopoulou, Hippokratio General Hospital Diabetes Centre; Athina Karabela, Dafni Psychiatric Hospital; Eirini Glezou and Marios Mangioros, Dromokaiteio Psychiatric Hospital; Angela Rentari, Harokopio University of Athens; and Danielle Walker, Wellcome Trust Sanger Institute. GOMAP-TEENAGE thanks the Sample Management and Genotyping Facilities staff at the Wellcome Trust Sanger Institute for sample preparation, quality control and genotyping.

Genomic Research Cohort for CCMB Diabetes Study (GRCCDS) comprises of various cohorts that are supported by: Council of Scientific Industrial Research (CSIR); Ministry of Science and Technology, Govt. of India, India; and Wellcome Trust, London, UK. GRCCDS is grateful to the patients and subjects who voluntarily participated in the study, and thankfully acknowledge other researchers who have supported the study.

Health, Aging and Body Composition Study (HABC) was supported by NIA contracts N01AG62101, N01AG62103, and N01AG62106. The genome-wide association study was funded by NIA grant 1R01AG032098-01A1 to Wake Forest University Health Sciences and genotyping services were provided by the Center for Inherited Disease Research (CIDR). CIDR is fully funded through a federal contract from the National Institutes of Health to The Johns Hopkins University, contract number HHSN268200782096C. This research was supported in part by the Intramural Research Program of the NIH, National Institute on Aging.

Healthy Aging in Neighborhoods of Diversity Across the Life Span Study (HANDLS) was supported by the Intramural Research Program of the NIH, National Institute on Aging (project Z01-AG000513 and human subjects' protocol 09 AGN248). Data analyses for HANDLS utilized the high-performance computational resources of the Biowulf Linux cluster at the National Institutes of Health, Bethesda, MD (http://hpc.nih.gov).

Hispanic Community Health Study/Study of Latinos (HCHS/SOL) is a collaborative study supported by contracts from the National Heart, Lung, and Blood Institute (NHLBI) to the University of North Carolina (HHSN268201300001I / N01-HC-65233), University of Miami (HHSN268201300004I / N01-HC-65234), Albert Einstein College of Medicine (HHSN268201300002I / N01-HC-65235), University of Illinois at Chicago (HHSN268201300003I / N01-HC-65236 Northwestern Univ), and San Diego State University (HHSN268201300005I / N01-HC-65237). The following Institutes/Centers/Offices have contributed to the HCHS/SOL through a transfer of funds to the NHLBI: National Institute on Minority Health and Health Disparities, National Institute on Deafness and Other Communication Disorders, National Institute of Dental and Craniofacial Research, National Institute of Diabetes and Digestive and Kidney Diseases, National Institute of Neurological Disorders and Stroke, NIH Institution-Office of Dietary Supplements. The Genetic Analysis Center at the University of Washington was supported by NHLBI and NIDCR contracts (HHSN268201300005C AM03 and MOD03).

Health Professionals' Follow-Up Study (HPFS) and Nurses Health Study (NHS) acknowledge assistance with data cleaning that was provided by the National Center for Biotechnology Information. Support for collection of datasets and samples was provided by the Collaborative Study on the Genetics of Alcoholism (COGA; U10 AA008401), the Collaborative Genetic Study of Nicotine Dependence (COGEND; P01 CA089392), and the Family Study of Cocaine Dependence (FSCD; R01 DA013423). Funding support for genotyping, which was performed at the Johns Hopkins University Center for Inherited Disease Research, was provided by the NIH GEI (U01HG004438), the National Institute on Alcohol Abuse and Alcoholism, the National Institute on Drug Abuse, and the NIH contract "High throughput genotyping for studying the genetic contributions to human disease" (HHSN268200782096C). The datasets used for the analyses described in this manuscript were obtained from dbGaP at http://www.ncbi.nlm.nih.gov/projects/gap/cgi-bin/study.cgi?study_id=phs000091.v1.p1 through dbGaP accession number phs000091.v1.p. Mexican American Hypertension and Insulin Resistance (HTNIR) was supported by grant HL0597974.

Hypertension and Insulin Resistance (HTN-IR) was supported by R01-HL067974, R01-HL-55005 and R01-HL 067974.

Howard University Family Study (HUFS) was supported by National Institutes of Health grants S06GM008016-320107 to CNR and S06GM008016-380111 to AA. Participant enrollment was carried out at the Howard University General Clinical Research Center, supported by National Institutes of Health grant 2M01RR010284. Genotyping support was provided by the Coriell Institute for Medical Research. This research was supported by the Intramural Research Program of the Center for Research on Genomics and Global Health (CRGGH). The CRGGH is supported by the National Human Genome Research Institute, the National Institute of Diabetes and Digestive and Kidney Diseases, the Center for Information Technology, and the Office of the Director at the National Institutes of Health (Z01HG200362).

INTERHEART (INTERHEART) was funded by: the Canadian Institutes of Health Research, the Heart and Stroke Foundation of Ontario, and the International Clinical Epidemiology Network (INCLEN); unrestricted grants from several pharmaceutical companies (with major contributions from AstraZeneca, Novartis, Hoechst Marion Roussel [now Aventis], Knoll Pharmaceuticals [now Abbott], Bristol-Myers Squibb, King Pharma, and Sanofi-Synthelabo); and various national bodies in different countries (see Online Appendix at http://image.thelancet.com/extras/04art8001webappendix2.pdf). Funding sources had no involvement in the study design; in the collection, analysis, and interpretation of data; or the writing of the manuscript.

The Jackson Heart Study (JHS) is supported by Contracts HHSN268201800010I, HHSN268201800011I, HHSN268201800012I, HHSN268201800013I, HHSN268201800014I, HHSN268201800015I from the National Heart, Lung, and Blood Institute (NHLBI) with additional support from the National Institute on Minority Health and Health Disparities (NIMHD). This manuscript has been reviewed by JHS for scientific content.

The views expressed in this manuscript are those of the authors and do not necessarily represent the views of the National Heart, Lung, and Blood Institute; the National Institutes of Health; or the U.S. Department of Health and Human Services.

Korean Association Resource (KARE) was supported by grants from Korea Centers for Disease Control and Prevention (4845–301, 4851–302, 4851–307) and intramural grants from the Korea National Institute of Health (2016-NI73001-00, 2019-NG-053-00). KARE was performed with bioresources from National Biobank of Korea, the Centers for Disease Control and Prevention, Republic of Korea.

Korea Biobank Array (KBA) Project was supported by an intramural grant from the National Institute of Health, Disease Control Prevention and Control Agency, Republic of Korea (2025-NI-002-00).

Korean Biobank Array from the Korean Genome and Epidemiology (KoGES) Consortium (KBA) was supported by grants from Korea Centers for Disease Control and Prevention (4845–301, 4851–302, 4851–307) and intramural grants from the Korea National Institute of Health (2016-NI73001-00, 2019-NG-053-00). KBA was performed with bioresources from National Biobank of Korea, the Centers for Disease Control and Prevention, Republic of Korea. Genotype data were provided by the Collaborative Genome Program for Fostering New Post-Genome Industry (3000-3031b).

Collaborative Health Research in the Region of Augsburg (KORA) research platform was initiated and financed by the Helmholtz Zentrum München – German Research Center for Environmental Health, which is funded by the German Federal Ministry of Education and Research and by the State of Bavaria. Furthermore, KORA research was supported within the Munich Center of Health Sciences (MC Health), Ludwig-Maximilians-Universität, as part of LMUinnovativ and by the German Center for Diabetes Research (DZD).

Los Angeles Latino Eye Study (LALES) acknowledges funding from NEI grant U10EY011753.

London Life Sciences Prospective Population (LOLIPOP) is supported by the National Institute for Health Research (NIHR) Comprehensive Biomedical Research Centre Imperial College Healthcare NHS Trust, the British Heart Foundation (SP/04/002), the MedicalResearch Council (G0601966, G0700931), the Wellcome Trust (084723/Z/08/Z, 090532 & 098381) the NIHR (RP-PG-0407-10371), the NIHR Official Development Assistance (ODA, award 16/136/68), the European Union FP7 (EpiMigrant, 279143) and H2020 programs (iHealth-T2D, 643774). LOLIPOP acknowledges support of the MRC-PHE Centre for Environment and Health, and the NIHR Health Protection Research Unit on Health Impact of Environmental Hazards. The work was carried out in part at the NIHR/Wellcome Trust Imperial Clinical Research Facility. The views expressed are those of the author(s) and not necessarily those of the Imperial College Healthcare NHS Trust, the NHS, the NIHR or the Department of Health. LOLIPOP thanks the participants and research staff who made the study possible.

Mexican American Study of Coronary Artery Disease (MACAD) was supported by grant R01-HL088457 and R01-HL-60030.

Mexico City T2D study (MC). In Mexico, this work was supported by the Fondo Sectorial de Investigación en Salud y Seguridad Social (SSA/IMSS/ISSSTE-CONACYT) project 150352, Temas Prioritarios de Salud Instituto Mexicano del Seguro Social 2014-FIS/IMSS/PROT/PRIO/14/34 and the Fundación IMSS. We thank Jorge Gutierrez Cuevas, Jaime Gómez Zamudio and Araceli Méndez Padrón for technical support. In Canada, the research was supported by a Canadian Institutes of Health Research (CIHR) operating grant to EJP and also by funding from the Banting and Best Diabetes Centre to EJP. In Canada, computations were performed on the GPC supercomputer at the SciNet HPC Consortium (https://scinethpc.ca/). SciNet is funded by Innovation, Science and Economic Development Canada; the Digital Research Alliance of Canada (https://alliancecan.ca/) ; the Ontario Research Fund: Research Excellence; and the University of Toronto.

Multi-Ethnic Study of Atherosclerosis (MESA). MESA and the MESA SHARe project are conducted and supported by the National Heart, Lung, and Blood Institue (NHLBI) in collaboration with MESA investigators. Support for MESA is provided by contracts 75N92025D00022, 75N92020D00001, HHSN268201500003I, N01-HC-95159, 75N92025D00026, 75N92020D00005, N01-HC-95160, 75N92020D00002, N01-HC-95161, 75N92025D00024, 75N92020D00003, N01-HC-95162, 75N92025D00027, 75N92020D00006, N01-HC-95163, 75N92025D00025, 75N92020D00004, N01-HC-95164, 75N92025D00028, 75N92020D00007, N01-HC-95165, N01-HC-95166, N01-HC-95167, N01-HC-95168, N01-HC-95169, UL1-TR-000040, UL1-TR-001079, UL1-TR-001420, UL1TR001881, DK063491, and R01HL105756. The authors thank the MESA participants and the MESA investigators and staff for their valuable contributions. A full list of participating MESA investigators and institutions can be found at http://www.mesa-nhlbi.org. This research was also supported by the Mexican-American Coronary Artery Disease (MACAD) National Heart, Lung, and Blood Institute, contracts R01-HL088457, R01-HL-60030; Hypertension and Insulin Resistance (HTN-IR) contracts R01-HL067974, R01-HL-55005, R01-HL 067974, and the Genetics of Latinos Diabetic Retinopathy (GOLDR) Study grant EY14684.

Metabolic Syndrome in Men (METSIM) was supported by the Academy of Finland (contract 124243), the Finnish Heart Foundation, the Finnish Diabetes Foundation, Tekes (contract 1510/31/06), and the Commission of the European Community (HEALTH-F2-2007 201681), and the US National Institutes of Health grants DK093757, DK072193, DK062370, and ZIA-HG000024.

Mexican Biobank (MXBB) was supported by Mexico's CONACYT (Grant number FONCICYT/50/2016; PI Moreno-Estrada), and the Newton Fund through the UK Medical Research Council (Grant number MR/N028937/1; PI Moreno-Estrada) to genetically characterize the population-based cohort derived from the National Health Survey 2000 (ENSA2000). The resulting ENSA Genomics Consortium acknowledges the seminal effort of Dr. Jaime Sepúlveda, the Mexican Ministry of Health, and the National Institute of Public Health, in the design and implementation of the ENSA2000 survey from which genomic data were generated for the MXB Project.

Mass General Brigham Biobank (MGBB) acknowledges the Partners HealthCare System for support of the MGB biobank and MGB patients for providing samples, genomic data, and health information data, as well as research support by NIDDK K24 DK110550 (to J.C.F.), K24 DK080140 (to J.B.M.) and NIDDK K23DK114551 (to M.S.U).

Michigan Genomics Initiative (MGI) was supported by NIH research grants HL117626 and HG007022. MGI was supported by internal research funds from the University of Michigan School of Public Health, the University of Michigan Medical School, and the University of Michigan President's Office. MGI are especially grateful to the generosity of all research participants.

VA Million Veteran Program (MVP). We gratefully acknowledge the Veterans who participated in the Million Veteran Program. This research is based on data from the Million Veteran Program, Office of Research and Development, and Veterans Health Administration (MVP003/028) as well as MVP000. This publication does not represent the views of the Department of Veterans Affairs or the United States Government. Support provided by BX003362 (Chang/Tsao), CSP2012 (JAL), R01DK134575 (MV). JAL and MV are also supported by MVP003.

The NAGAHAMA Study is a community-based longitudinal study in Japan. Participants in the Nagahama study were recruited from general population of Nagahama, a rural city of 125,000 inhabitants located in central Japan (N = 9,764). Community residents, aged between 30 and 74 years at recruitment, who were living independently without physical impairment or dysfunction, were eligible. All study procedures were approved by the Ethics Committee of Kyoto University Graduate School of Medicine and the Nagahama Municipal Review Board. Written informed consent was obtained from all participants.

Netherlands Epidemiology of Obesity (NEO) thanks all individuals who participated in the study, all participating general practitioners for inviting eligible participants and all research nurses for collection of the data. NEO thank the study group, Pat van Beelen, Petra Noordijk and Ingeborg de Jonge for the coordination, lab and data management of the study. Genotyping was supported by the Centre National de Génotypage (Paris, France), headed by Jean-Francois Deleuze. NEO is supported by the participating Departments, the Division and the Board of Directors of the Leiden University Medical Center, and by the Leiden University, Research Profile Area Vascular and Regenerative Medicine. NIDDM-Atherosclerosis Study Hispanic Cohorts (NIDDM) was supported by grant HL055798.

Northwestern University Genetics (NUGENE) was funded by the Northwestern University's Center for Genetic Medicine, Northwestern University, and Northwestern Memorial Hospital. Samples and data used in this study were provided by the NUgeneProject (www.nugene.org). Assistance with phenotype harmonization was provided by the eMERGE Coordinating Center (Grant number U01HG04603). This study was funded through the NIH, NHGRI eMERGE Network (U01HG004609). Funding support for genotyping, which was performed at The Broad Institute, was provided by the NIH (U01HG004424). Assistance with phenotype harmonization and genotype data cleaning was provided by the eMERGE Administrative Coordinating Center (U01HG004603) and the National Center for Biotechnology Information (NCBI). The datasets used for the analyses described in this manuscript were obtained from dbGaP at http://www.ncbi.nlm.nih.gov/gap through dbGaP accession number phs000237.v1.p1.

Prospective Investigation of the Vasculature in Uppsala Seniors (PIVUS) was supported by Wellcome Trust Grants (WT098017, WT064890, WT090532), Uppsala University, Uppsala University Hospital, the Swedish Research Council, and the Swedish Heart-Lung Foundation. Penn Medicine BioBank (PMBB). We acknowledge the PMBB for providing data and thank the patient-participants of Penn Medicine who consented to participate in this research program. We would also like to thank the Penn Medicine BioBank team and Regeneron Genetics Center for providing genetic variant data for analysis. The PMBB is approved under IRB protocol# 813913 and supported by Perelman School of Medicine at University of Pennsylvania, a gift from the Smilow family, and the National Center for Advancing Translational Sciences of the National Institutes of Health under CTSA award number UL1TR001878.

Pakistan Risk of Myocardial Infarction Study (PROMIS) was funded by the Wellcome Trust, UK, and Pfizer (genotyping) and was supported through funds available to investigators at the Center for Non-Communicable Diseases, Pakistan, and the University of Cambridge, UK (fieldwork). Biomarker assays in PROMIS have been funded through grants awarded by the National Institutes of Health (RC2HL101834 and RC1TW008485) and the Fogarty International (RC1TW008485).

Prospective Study of Pravastatin in the Elderly at Risk (PROSPER) was supported by an investigator-initiated grant obtained from Bristol-Myers Squibb. Prof. J.W.J. is an Established Clinical Investigator of the Netherlands Heart Foundation (grant 2001 D 032). Support for genotyping was provided by the seventh framework program of the European commission (grant 223004) and by the Netherlands Genomics Initiative (Netherlands Consortium for Healthy Aging grant 050-060-810).

Sea Islands Genetic Network Reasons for Geographic and Racial Differences in Stroke (REGARDS) is supported by cooperative agreement U01 NS041588 co-funded by the National Institute of Neurological Disorders and Stroke (NINDS) and the National Institute on Aging (NIA), National Institutes of Health, Department of Health and Human Service. The content is solely the responsibility of the authors and does not necessarily represent the official views of the NINDS or the NIA. Additional funding was from R01 DK084350 from the National Institutes of Health.

Ragama Health Study (RHS) was supported by a grant from the National Center for Global Health and Medicine (NCGM)

Rotterdam Study (RS) is grateful to the participants and staff involved in the study, and the participating general practitioners and pharmacists. RS is funded by Erasmus Medical Center and Erasmus University, Rotterdam, Netherlands Organization for the Health Research and Development (ZonMw), the Research Institute for Diseases in the Elderly (RIDE), the Ministry of Education, Culture and Science, the Ministry for Health, Welfare and Sports, the European Commission (DG XII), and the Municipality of Rotterdam.

San Antonio Family Heart Study (SAFHS) was supported by U01 DK085524, R01 HL113323, P01 HL045222, R01 DK047482, and R01 DK053889. The Veterans Administration Genetic Epidemiology Study (VAGES) study was supported by a Veterans Administration Epidemiologic grant. The Family Investigation of Nephropathy and Diabetes - San Antonio (FIND-SA) study was supported by NIH grant U01 DK57295. The SAMAFS research team acknowledges late Dr. Hanna E. Abboud's contributions to the research activities of the SAMAFS.

Shanghai Breast Cancer Study and Shanghai Women's Health Study (SBCS/SWHS) was supported in part by US National Institutes of Health grants R01CA64277 and R01CA124558, as well as Ingram Professorship and Research Reward funds from the Vanderbilt University School of Medicine. We want to thank participants and research staff of the study, Regina Courtney for plasma and DNA sample preparation, and Hui Cai, Ben Zhang and Jing He for data processing and analyses.

Singapore Chinese Eye Study (SCES) is supported by the National Medical Research Council (NMRC), Singapore (grants 0796/2003, 1176/2008, 1149/2008, STaR/0003/2008, 1249/2010, CG/SERI/2010, CIRG/1371/2013, and CIRG/1417/2015), and Biomedical Research Council (BMRC), Singapore (08/1/35/19/550 and 09/1/35/19/616).

Starr County Health (SCH) was supported by grants from the National Institutes of Health (DK073541, DK085501, HL102830 and DK116378) and funds from the State of Texas. SCH thank the field staff in Starr County for their careful collection of these data and are especially grateful to the participants who so graciously cooperated and gave of their time. Starr County Health Singapore Chinese Health Study (SCHS) was supported by the US National Institutes of Health grants R01DK08072, R01CA144034 and UM1CA182876.

Slim Initiative for Genomic Medicine in the Americas (SIGMA). This work was conducted as part of the Slim Initiative for Genomic Medicine, a joint U.S.-Mexico project funded by the Carlos Slim Health Institute. The UNAM/INCMNSZ diabetes study was supported by Consejo Nacional de Ciencia y Tecnología grants 138826, 128877, CONACyT- SALUD 2009-01-115250, and a grant from Dirección General de Asuntos del Personal Académico, UNAM, IT 214711. The Diabetes in Mexico Study was supported by Consejo Nacional de Ciencia y Tecnología grant 86867 and by Instituto Carlos Slim de la Salud, A.C. The Mexico City Diabetes Study was supported by National Institutes of Health (NIH) grant R01HL24799 and by the Consejo Nacional de Ciencia y Tenologia grants: 2092, M9303, F677-M9407, 251M, and 2005-C01-14502, SALUD 2010-2-151165. The Multiethnic Cohort was supported by NIH grants CA164973, CA054281, and CA063464.

Singapore Malay Eye Study (SIMES) is supported by the National Medical Research Council (NMRC), Singapore (grants 0796/2003, 1176/2008, 1149/2008, STaR/0003/2008, 1249/2010, CG/SERI/2010, CIRG/1371/2013, and CIRG/1417/2015), and Biomedical Research Council (BMRC), Singapore (08/1/35/19/550 and 09/1/35/19/616).

Singapore Indian Eye Study (SINDI) is supported by the National Medical Research Council (NMRC), Singapore (grants 0796/2003, 1176/2008, 1149/2008, STaR/0003/2008, 1249/2010, CG/SERI/2010, CIRG/1371/2013, and CIRG/1417/2015), and Biomedical Research Council (BMRC), Singapore (08/1/35/19/550 and 09/1/35/19/616).

Samsung Medical Center (SMC) was supported by a grant from Samsung Biomedical Research Institute. Genotyping of the patients and control subjects from SMC was conducted by Duk-Hwan Kim in the Dept. of Molecular Cell Biology, Sungkyunkwan University School of Medicine, and was supported by a grant from Samsung Biomedical Research Institute.

Seoul National University Hospital (SNUH) was supported by a grant from the Korea Health Technology R&D Project through the Korea Health Industry Development Institute, funded by the Ministry of Health & Welfare (grant numbers HI15C1595, HI14C0060, HI15C3131).

Taiwan Metabochip Consortium Zhonghua (TAICHI-G) was supported by grants from: the National Health Research Institutes, Taiwan (PH-099-PP-03, PH-100-PP-03, and PH-101-PP- 03); the National Science Council, Taiwan (NSC 101-2314-B-075A-006-MY3, MOST 104-2314- B-075A-006-MY3, MOST 104-2314-B-075A-007, and MOST 105-2314-B-075A-003); and the Taichung Veterans General Hospital, Taiwan (TCVGH-1020101C, TCVGH-1020102D, TCVGH-1023102B, TCVGH-1023107D, TCVGH-1030101C, TCVGH- 1030105D, TCVGH-1033503C, TCVGH-1033102B, TCVGH-1033108D, TCVGH-1040101C, TCVGH-1040102D, TCVGH-1043504C, and TCVGH-1043104B). TAICHI-G was also supported in part by the National Center for Advancing Translational Sciences (CTSI grant UL1TR001881).

Taiwan Type 2 Diabetes (TWT2D) was supported by the GMM Study, Academia Sinica, Taiwan.

Danish T2D Case-Control Study (UCPH) was undertaken by the Novo Nordisk Foundation Center for Basic Metabolic Research, which is an independent Research Center, based at the University of Copenhagen, Denmark and partially funded by an unconditional donation from the Novo Nordisk Foundation (www.cbmr.ku.dk, Grant number NNF18CC0034900). Included study samples were supported by the Danish Research Fund and the National Danish Research Fund (The Vejle Diabetes Biobank), the Velux Foundation, The Danish Medical Research Council and Danish Agency for Science, Technology and Innovation (Health 2006); the Danish Research Council, the Danish Centre for Health Technology Assessment and Novo Nordisk Inc. (Inter99), the Timber Merchant Vilhelm Bang's Foundation and the Danish Heart Foundation (Health 2008), TrygFonden, the Lundbeck Foundation and the Novo Nordisk Foundation (NNF15OC0015896, DanFunD).

UK Biobank (UKBB) analyses were conducted using the UK Biobank resource under applications 236, 9161, and 10035. This research was supported by the British Heart Foundation (grant SP/13/2/30111). Large-scale comprehensive genotyping of UK Biobank for cardiometabolic traits and diseases: UK CardioMetabolic Consortium (UKCMC).

Uppsala Longitudinal Study of Adult Men (ULSAM) was supported by Wellcome Trust Grants (WT098017, WT064890, WT090532), Uppsala University, Uppsala University Hospital, the Swedish Research Council, and the Swedish Heart-Lung Foundation.

Wake Forest School of Medicine (WFSM) was supported by NIH grants K99 DK081350, R01 DK066358, R01 DK053591, R01 DK087914, U01 DK105556, R01 HL56266, R01 DK070941 and in part by the General Clinical Research Center of the Wake Forest School of Medicine grant M01 RR07122. Genotyping services were provided by the Center for Inherited Disease Research (CIDR), which is fully funded through a federal contract from the National Institutes of Health to The Johns Hopkins University, contract number HHSC268200782096C.

Women's Health Initiative (WHI). The WHI program is funded by the National Heart, Lung, and Blood Institute, National Institutes of Health, U.S. Department of Health and Human Services through contracts 75N92021D00001, 75N92021D00002, 75N92021D00003, 75N92021D00004, 75N92021D00005.

Wellcome Trust Case Control Consortium (WTCCC) analysis and genotyping was supported by: Wellcome Trust funding 090367, 098381, 090532, 083948, 085475, 101630, and 203141; MRC (G0601261); EU (Framework 7) HEALTH-F4-2007-201413; and NIDDK DK098032 and U01-DK105535.

**Genes & Health Research Team authorship for Scientific Publications**

| | | |
|---|---|---|
| Eamonn Maher | | Aston University |
| Shabana Chaudhary | | Blizard Institute, Queen Mary University of London |
| Joseph Gafton | | Blizard Institute, Queen Mary University of London |
| Karen A Hunt | | Blizard Institute, Queen Mary University of London |
| Shapna Hussain | | Blizard Institute, Queen Mary University of London |
| Kamrul Islam | | Blizard Institute, Queen Mary University of London |
| Mohammed Bodrul Mazid | | Blizard Institute, Queen Mary University of London |
| Elizabeth Owor | | Blizard Institute, Queen Mary University of London |
| Jessry Russell | | Blizard Institute, Queen Mary University of London |
| Nishat Safa | | Blizard Institute, Queen Mary University of London |
| John Solly | | Blizard Institute, Queen Mary University of London |
| Marie Spreckley | | Blizard Institute, Queen Mary University of London |
| David A Van Heel | | Blizard Institute, Queen Mary University of London |
| Jan Whalley | | Blizard Institute, Queen Mary University of London |
| Ishevanhu Zengeya | | Blizard Institute, Queen Mary University of London |
| Emily Mantle | | Blizard Institute, Queen Mary University of London |
| Shaheen Akhtar | | Bradford Teaching Hospitals NHS Foundation Trust |
| Samina Ashraf | | Bradford Teaching Hospitals NHS Foundation Trust |
| Dan Mason | | Bradford Teaching Hospitals NHS Foundation Trust |

John Wright Bradford Teaching Hospitals NHS Foundation Trust
Daniel MacArthur Garvan Institute
Michael Simpson King's College London
Richard C Trembath King's College London
Gerome Breen Kings College London
Raymond Chung College London
Sang Hyuck Lee College London
Omar Asgar Manchester University Hospitals
Joanne Harvey Manchester University Hospitals
Karen Tricker Manchester University Hospitals
Caroline Winckley Manchester University Hospitals
Hanifa Khatun Manchester University Hospitals
Amna Asif Manchester University Hospitals
Claudia Langenberg Precision Healthcare University Research Institute, Queen Mary University of London
Grainne Colligan Social Action for Health (charity)
Ceri Durham Social Action for Health (charity)
Bill Newman University of Manchester
Ahsan Khan Waltham Forest Council
Hilary Martin Wellcome Sanger Institute
Teng Heng Wellcome Sanger Institute
Matt Hurles Wellcome Sanger Institute
Vivek Iyer Sanger Institute
Georgios Kalantzis Wellcome Sanger Institute
Vladimir Ovchinnikov Wellcome Sanger Institute
Iaroslav Popov Wellcome Sanger Institute
Klaudia Walter Wellcome Sanger Institute
Panos Deloukas William Harvey Research Institute, Queen Mary University of London
David Collier William Harvey Research Institute, Queen Mary Universityof London
Ana Angel Wolfson Institute of Population Health, Queen Mary University of London
Saeed Bidi Wolfson Institute of Population Health, Queen Mary University of London
Fabiola Eto Institute of Population Health, Queen Mary University of London
Sarah Finer Wolfson Institute of Population Health, Queen Mary University of London
Chris Griffiths Wolfson Institute of Population Health, Queen Mary University of London
Sam Hodgson Wolfson Institute of Population Health, Queen Mary University of London
Benjamin M Jacobs Wolfson Institute of Population Health, Queen Mary University of London
Rohini Mathur Wolfson Institute of Population Health, Queen Mary University of London
Caroline Morton Wolfson Institute of Population Health, Queen Mary University of London
Asma Qureshi Wolfson Institute of Population Health, Queen Mary University of London
Stuart Rison Wolfson Institute of Population Health, Queen Mary University of London
Annum Salman Wolfson Institute of Population Health, Queen Mary University of London
Miriam Samuel Wolfson Institute of Population Health, Queen Mary University of London
Moneeza K Siddiqui Wolfson Institute of Population Health, Queen Mary University of London

| | | |
|---|---|---|
| Daniel Stow | | Wolfson Institute of Population Health, Queen Mary University of London |
| Sabina Yasmin | | Wolfson Institute of Population Health, Queen Mary University of London |
| Julia Zöllner | | Wolfson Institute of Population Health, Queen Mary University of London |
| Sheik Dowlut | | Wolfson Institute of Population Health, Queen Mary University of London |
